# Polygenic risk scores validated in patient-derived cells stratify for mitochondrial subtypes of Parkinson’s disease

**DOI:** 10.1101/2023.05.12.23289877

**Authors:** Giuseppe Arena, Zied Landoulsi, Dajana Grossmann, Armelle Vitali, Sylvie Delcambre, Alexandre Baron, Paul Antony, Ibrahim Boussaad, Dheeraj Reddy Bobbili, Ashwin Ashok Kumar Sreelatha, Lukas Pavelka, Christine Klein, Philip Seibler, Enrico Glaab, Manu Sharma, Rejko Krüger, Patrick May, Anne Grünewald the NCER-PD and COURAGE-PD Consortia

**Affiliations:** Luxembourg Centre for Systems Biomedicine, University of Luxembourg; L-4367, Esch-sur-Alzette, Luxembourg; Translational Neurodegeneration Section “Albrecht-Kossel”, Department of Neurology, University Medical Center Rostock, University of Rostock; D-18147, Rostock, Germany; Centre for Genetic Epidemiology, Institute for Clinical Epidemiology and Applied Biometry, University of Tübingen; D-72076, Tübingen, Germany; Transversal Translational Medicine, Luxembourg Institute of Health; L-1445, Strassen, Luxembourg; Centre Hospitalier du Luxembourg, Parkinson Research Clinic; L-1210, Luxembourg, Luxembourg; Institute of Neurogenetics, University of Lübeck; D-23538, Lübeck, Germany

**Keywords:** idiopathic Parkinson’s disease, mitochondrial common variants, polygenic risk scores, oxidative phosphorylation, functional validation

## Abstract

**Background:** Parkinson’s disease (PD) is the fastest growing neurodegenerative disorder, with affected individuals expected to double during the next 20 years. This raises the urgent need to better understand the genetic architecture and downstream cellular alterations underlying PD pathogenesis, in order to identify more focused therapeutic targets. While only ∼10% of PD cases can be clearly attributed to monogenic causes, there is mounting evidence that additional genetic factors could play a role in idiopathic PD (iPD). In particular, common variants with low to moderate effect size in multiple genes regulating key neuroprotective activities may act as risk factors for PD. In light of the well-established involvement of mitochondrial dysfunction in PD, we hypothesized that a fraction of iPD cases may harbour a pathogenic combination of common variants in nuclear-encoded mitochondrial genes, ultimately resulting in neurodegeneration.

**Methods:** to capture this mitochondria-related “missing heritability”, we leveraged on existing data from previous genome-wide association studies (GWAS) – i.e., the large PD GWAS from Nalls and colleagues. We then used computational approaches based on mitochondria-specific polygenic risk scores (mitoPRSs) for imputing the genotype data obtained from different iPD case-control datasets worldwide, including the Luxembourg Parkinson’s Study (412 iPD patients and 576 healthy controls) and the COURAGE-PD cohorts (7270 iPD cases and 6819 healthy controls).

**Results:** applying this approach to gene sets controlling mitochondrial pathways potentially relevant for neurodegeneration in PD, we demonstrated that common variants in genes regulating *Oxidative Phosphorylation (OXPHOS*-PRS) were significantly associated with a higher PD risk both in the Luxembourg Parkinson’s Study (odds ratio, OR=1.31[1.14-1.50], *p*=5.4e-04) and in COURAGE-PD (OR=1.23[1.18-1.27], *p*=1.5e-29). Functional analyses in primary skin fibroblasts and in the corresponding induced pluripotent stem cells-derived neuronal progenitor cells from Luxembourg Parkinson’s Study iPD patients stratified according to the *OXPHOS*-PRS, revealed significant differences in mitochondrial respiration between high and low risk groups (*p* < 0.05). Finally, we also demonstrated that iPD patients with high *OXPHOS*-PRS have a significantly earlier age at disease onset compared to low-risk patients.

**Conclusions:** our findings suggest that OXPHOS-PRS may represent a promising strategy to stratify iPD patients into pathogenic subgroups – in which the underlying neurodegeneration is due to a genetically defined mitochondrial burden – potentially eligible for future, more tailored mitochondrially targeted treatments.

## Background

Parkinson’s disease (PD) is one of the commonest neurodegenerative disorders, affecting about 1-2% of the population over the age of 60 [1]. At clinical presentation, PD patients typically display bradykinesia with rigidity and/or resting tremor accompanied by a heterogeneous panel of non-motor symptoms (e.g., sleep disturbance, mood disorders, cognitive changes, and autonomic dysfunction). Motor symptoms are mostly caused by disrupted dopamine signaling in the striatum because of dopaminergic neuron (DAN) loss in the *substantia nigra pars compacta* of the brain. At the same time, typical proteinaceous inclusions (i.e., Lewy bodies), composed of α-synuclein, ubiquitin and other aggregated proteins or organelles, are observed in the surviving DANs [2]. The etiology of PD is complex and influenced by both environmental and genetic factors. Monogenic familial forms, clearly ascribable to single mutations in autosomal dominant (e.g., *SNCA*, *LRRK2*, *VPS35*) or recessive (*PRKN*, *PINK1*, *PARK7*) genes, account for approximately 5-10% of all PD cases [3]. However, the contribution of genetics in the remaining 90-95% of patients with idiopathic PD (iPD) remains poorly understood, and disease susceptibility may be significantly influenced by the synergistic effect of multiple common low-risk genetic variants [4]. To decipher this “missing heritability”, systematic approaches based on polygenic risk scores (PRSs) have been proposed in recent years, aimed at identifying individuals with a higher risk to develop PD or exhibiting specific clinical phenotypes [5]. PRSs are calculated as the sum of common risk single nucleotide polymorphisms (SNPs) weighted by their genome-wide associated studies (GWAS) effect sizes [6]. Applying PRS to a restricted number of genes regulating biological functions known to be altered in PD could potentially reveal a relationship between the pathways involved and their impact in determining the clinical phenotype. Such approach would allow the stratification of patients according to the underlying pathological mechanisms, thus enabling precision medicine therapeutic approaches [5].

There is convincing and longstanding evidence that points to mitochondrial dysfunction as an early and causative event in PD pathogenesis. Indeed, both epidemiological studies on humans exposed to pesticides as well as toxin-induced PD models support a primary role for impaired mitochondrial electron transport chain (ETC) activity, as suggested by the selective degeneration of DANs following mitochondrial complex I disruption [7,8]. Moreover, molecular genetic studies in PD families revealed that proteins mutated in early-onset forms (i.e., PINK1 and Parkin) regulate mitochondrial function or even localize to mitochondria [9]. Hallmarks of mitochondrial dysfunction have also been observed in cellular models established from iPD patients, showing defective oxidative phosphorylation (OXPHOS), increased oxidative stress, and mitochondrial DNA (mtDNA) damage [10–12]. Neurodegeneration in such iPD patients cannot be explained by ageing or environmental factors alone [13], which possibly implies the existence of pathogenic variants in mitochondrial genes. In accordance with this possibility, a recent study on mitochondria-specific PRSs (mitoPRSs) demonstrated that combinations of small effect variants within genes regulating mitochondrial function were statistically associated with higher PD risk [14].

Herein, we extended the mitoPRS concept to subsets of genes controlling distinct mitochondrial pathways and demonstrated, in two independent patient datasets from large and deep phenotyped PD cohorts worldwide, that common variants in genes regulating *Oxidative Phosphorylation* (*OXPHOS*) were significantly associated with an increased risk of developing PD. Importantly, we functionally validated individual *OXPHOS*-PRS profiles in the corresponding patient-based cellular models, showing significant differences in mitochondrial respiratory function between iPD patients with high or low *OXPHOS*-PRS. Finally, available clinical data from the genetically and functionally stratified individuals revealed that iPD patients with high *OXPHOS*-PRSs displayed a significantly earlier age at disease onset compared to low *OXPHOS*-PRS patients.

## Methods

### Study design

Two datasets were analyzed for this work. First, the exploratory dataset from the ongoing Luxembourg Parkinson’s Study, a large longitudinal monocentric observational study in the framework of the NCER-PD (National Centre for Excellence in Research in Parkinson’s Disease) project, which aims at recruiting and following up patients with PD and other forms of neurodegenerative parkinsonism along with healthy controls (HC) [15]. At the time of data export, the Luxembourg Parkinson’s Study comprised 493 PD patients and 625 HC. Second, the replication dataset from the COURAGE-PD (Comprehensive Unbiased Risk Factor Assessment for Genetics and Environment in Parkinson’s Disease) consortium, including 21 sub-cohorts of European descent, but excluding the Luxembourg Parkinson’s Study and the International Parkinson’s Disease Genomics Consortium (IPDGC) samples (7422 PD patients and 6904 HC) [16,17].

### Clinical assessment

All participants in the Luxembourg Parkinson’s Study underwent a comprehensive clinical assessment, as described in detail elsewhere [15]. The diagnosis of PD was based on UKPDSBB diagnostic criteria [18]. For this study, we focused on eight PD-specific clinical outcomes, namely the Movement Disorder Society update of the Unified Parkinson’s Disease Rating Scale I (MDS-UPDRS I), MDS-UPDRS II, MDS-UPDRS III, MDS-UPDRS-IV, the Quality of Life questionnaire (PDQ39), the L-dopa-equivalent daily dose (LEDD), the Scales for Outcomes in Parkinson’s Disease – Autonomic Dysfunction (SCOPA-AUT) and the Montreal Cognitive Assessment (MoCA). Age at onset (AAO) was set as age at PD diagnosis. Disease duration corresponds to the duration in years since the official PD diagnosis to the point of data collection, i.e., until the baseline visits for all sample datasets analyzed.

### Genotyping and quality control

DNA samples of participants from both cohorts were genotyped using the Neurochip array (v.1.0 and v1.1; Illumina, San Diego, CA) that was specifically designed to integrate neurodegenerative disease-related variants [19]. Quality control (QC) of genotyping data was performed using PLINK v1.9 [20], as follows: samples with call rates < 95% and whose genetically determined sex deviated from gender reported in clinical data were excluded from the analysis, and the filtered variants were checked for cryptic relatedness and excess of heterozygosity. Samples exhibiting excess heterozygosity (F statistic > 0.2) and first-degree relatedness were excluded. Once sample QC was completed, SNPs with Hardy-Weinberg equilibrium P value < 1E-6, and missingness rates > 5% were excluded too. QC was conducted independently for each European cohort of the COURAGE-PD study, according to the standard procedures reported previously [17]. For both cohorts, we also excluded carriers of pathogenic PD-linked variants in eight PD-related genes (*ATP13A2*, *LRRK2*, *GBA*, *PARK7, PINK1, PRKN, VPS35* and *SNCA*) identified *via* genotyping data. For the Luxembourg Parkinson’s Study, the presence of these mutations was confirmed by Sanger sequencing. To consider the population stratification, we calculated the first three principal components (PCs) using PLINK. Genotyping data were then imputed using the Haplotype Reference Consortium r1.1 2016 of the Michigan Imputation Server and filtered for imputation quality (R2 > 0.3) [21].

### Mitochondrial gene sets and pathway resources

To assess the potential association between common variants in nuclear-encoded mitochondrial genes and PD risk, we first selected three different mitochondrial gene sets: Human MitoCarta3.0, which is a public inventory of 1136 genes encoding proteins with anticipated mitochondrial localization [22], and two gene sets previously reported by Billingsley and colleagues; the primary list (Billingsley I) contained 178 genes implicated in mitochondrial disorders, whereas the secondary list (Billingsley II) contained 1327 genes regulating, more generally, mitochondrial function [14].

Moreover, we defined six groups of genes – obtained from the Molecular Signatures Database (MsigDB) v7.5.1 – known to participate in the following mitochondrial pathways potentially related to PD pathogenesis: *Mitochondrial DNA regulation, Mitophagy, Oxidative phosphorylation, TCA cycle, Mitochondrial protein import* and *Mitochondrial proton transport* (**Supplementary Table 1**).

### Calculation of Polygenic Risk Scores (PRSs)

The genome-wide PD-PRSs were calculated using the PRSice2 R package with default settings [23]. PRSs for each individual were generated by summing the weighted effects of the risk alleles (including all variants at threshold below genome-wide significance) associated with PD – based on the largest PD GWAS summary statistics to date [4] – which are present in the two imputed target datasets (Luxembourg Parkinson’s study and COURAGE-PD genotype data). PRSs for the general or pathway-specific mitochondrial gene sets (mitoPRSs) were generated using the PRSet function in PRSice2, using only risk alleles within gene regions outlined in the different gene lists. PRSice2 implement the clumping and thresholding (C+T) method. The criteria for linkage disequilibrium (LD) clumping of SNPs were pairwise LD r2 < 0.1 within a 250 kb window. PRSs were computed at different P-value thresholds ranging from 5e-08 to 0.5. PRSice2 identified the optimum P-value threshold for variant selection that explains the maximum variance in the target sample. PRSice2 was also used to determine the observed phenotypic variance (PRS model fit, R2) explained by the genetic contribution of each mitochondrial pathways-PRS.

To control the specificity of the contribution of PD GWAS risk alleles in the prediction of PD, we calculated PRS for the Luxembourg Parkinson’s Study using SNPs from three base summary statistics of other disorders, namely Type 2 Diabetes, schizophrenia, and Alzheimer’s disease [24–26].

### Fibroblasts selection and culture

Skin biopsies from the lower back region of both iPD patients and HC showing the highest or lowest *OXPHOS*-PRSs (10th vs 90th percentile, n=4 for each group) were collected in low-glucose Dulbecco’s Modified Eagle Medium (DMEM, Thermo Fisher Scientific, #21885108) supplemented with 1% (v/v) Penicillin-Streptomycin (P/S, Thermo Fisher Scientific, #15140163). Each skin biopsy (5mm diameter punch) was cut into several (up to 5) pieces and placed with the dermis facing down into a cell culture flask containing heat-inactivated Fetal Bovine Serum (HI-FBS, Thermo Fisher Scientific, #10270-106). After 10 minutes incubation at room temperature, DMEM containing 4.5g/L Glucose and 4mM L-Glutamine (Thermo Fisher Scientific, #41965039), supplemented with 10% (v/v) HI-FBS and 1% (v/v) P/S, was added to the flask, followed by incubation at 37°C in a 5% CO_2_ humidified atmosphere. Cells were kept in the same medium for subsequent subculture, and subjected to Mycoplasma analysis on a regular basis to exclude potential contamination. The 16 fibroblasts lines all had the same passage number at the time of the experiments, ranging from passage 5 to 10.

### iPSC generation

Fibroblast reprogramming into induced pluripotent stem cells (iPSCs) was performed by using the CytoTune^TM^-iPS 2.0 Sendai Reprogramming kit (Thermo Fisher Scientific, #A16518), following the manufacturer’s instructions (feeder-free conditions). Briefly, fibroblasts were transduced overnight with the reprogramming vectors hKOS, hc-Myc and hKlf4, at MOI 5, 5 and 3 respectively, and then maintained in standard fibroblast medium until day 7 post-transduction. On day 7, fibroblasts under reprogramming were plated on Geltrex (Thermo Fisher Scientific, #A1413202)-coated wells, and switched to iPSCs medium starting from day 8. iPSCs medium consisted in DMEM-F12, Hepes (Thermo Fisher Scientific, #31330038), supplemented with 1% P/S, 1% Insulin-Transferrin-Selenium (ITS-G, Thermo Fisher Scientific, #41400045), 64 µG/mL L-Ascorbic acid 2-phosphate magnesium (Sigma, #A8960), 100ng/ml heparin (Sigma, #H3149-25KU), 2ng/ml TGF-β1 (Peprotech, #100-21) and 10ng/ml FGF-basic (Peprotech, #100-18B). Between day 21 and 28 post-transduction, emerging undifferentiated iPSC colonies were manually transferred into Geltrex-coated 12-well plates for further expansion and characterization.

### Generation and maintenance of neuronal progenitor cells

The procedure for obtaining neuronal progenitor cells from iPSCs was described previously. Their identity is restricted to both midbrain and hindbrain fates, whereas they are not able to form forebrain neurons [27]. Briefly, iPSCs were shifted to N2B27 medium – consisting of 49% DMEM/F12 (Thermo Fisher Scientific, #21331020), 49% Neurobasal (Thermo Fisher Scientific, #21103049), 1:100 B27 supplement without vitamin A (Thermo Fisher Scientific, #12587010), 1:200 N2 supplement (Thermo Fisher Scientific, #17502001), 1% (v/v) Glutamax (Thermo Fisher Scientific, #35050061) and 1% (v/v) P/S – supplemented with 10µM SB-431542 (R&D Systems, #1614/10), 1µM Dorsomorphin (R&D Systems, #3093), 3µM CHIR 99021 (CHIR, Axon Medchem, #2435) and 0.5µM purmorphamine (PMA, Sigma, #SML0868). After 4 days in this medium, SB-431542 and Dorsomorphin were removed and cells maintained in N2B27 medium containing CHIR, PMA and 150µM Ascorbic Acid (AA, Sigma, #A4403). On day 5, the emerging neural epithelium was isolated, triturated into smaller pieces and plated on Geltrex-coated wells. The resulting neuronal progenitor cells (smNPCs) were further expanded in N2B27 medium supplemented with CHIR, PMA and AA.

smNPCs from two PD patients carrying the c.1366C>T mutation in the *PINK1* gene (p.Q456X) were described previously [28].

### Analysis of mitochondrial respiration

Oxygen consumption rates (OCRs) were determined by using the Seahorse XFe96 FluxPak (Agilent, #102416-100) and XF Cell Mito Stress Test (Agilent, #103015-100) kits, in accordance with the manufacturer’s instructions. For primary skin fibroblasts, 5000 cells per well were seeded in a 96-well Seahorse cell culture plate (at least 5 replicates per fibroblast line) and incubated overnight. The following day, fibroblast medium in each well was replaced with 175µl Seahorse XF DMEM Medium pH 7.4 (Agilent, #103575-100), supplemented with 25mM Glucose, 2mM L-Glutamine and 1mM Sodium Pyruvate, followed by 1h incubation at 37°C without CO_2_. In the meantime, the sensor cartridge – previously equilibrated overnight in the XF Calibrant solution at 37°C without CO_2_ – was loaded with standard mitochondrial toxins, namely Oligomycin (1µM final concentration in the assay well), carbonyl cyanide p-trifluoromethoxyphenylhydrazone (FCCP, 0.6µM final concentration in the assay well) and a mixture of Rotenone and Antimycin A (both at the concentration of 0.5µM in the assay well). After an additional incubation at 37°C without CO_2_ for 30 minutes, the sensor cartridge, as well as the cell culture plate, were loaded on the XFe96 Extracellular flux Analyzer (Agilent). Three OCR measurements were performed for both basal respiration and after each automated injection of mitochondrial toxins. Once the run was completed, the cell culture medium was removed from each well and DNA content measured by using the CyQUANT cell proliferation assay kit (Thermo Fisher Scientific, #C7026). OCR data were analyzed using the XFe Wave software (Agilent) and normalized against cell number, as defined by the CyQUANT assay. Normalized data were finally exported using the Seahorse XF Cell Mito Stress Test Report Generator and statistical analysis performed on at least three independent biological replicates. Initial OCR values as well as OCR measurements after each injection step were used to calculate different parameters related to oxidative phosphorylation, including basal respiration, proton leakage across the inner mitochondrial membrane, respiration coupled to ATP synthesis, maximal respiration and spare respiratory capacity [29].

Seahorse experiments in neuronal progenitor cells were performed in Neurobasal medium containing either glucose or galactose as carbon source [30]. Briefly, 35000 cells per well were first seeded in a Geltrex-coated 96-well Seahorse cell culture plate (at least 10 replicates per line), in 100µl N2B27 medium containing CHIR, PMA and AA. The day after, N2B27 medium was removed and replaced by Neurobasal-A without glucose and sodium pyruvate (Thermo Fisher Scientific, #A2477501), then supplemented with 0.727mM sodium pyruvate and either 25mM Glucose or 25mM Galactose. Similar to the standard N2B27 used for maintenance of neuronal progenitor cells, this medium also contained N2, B27 and Glutamax supplements, 1% (v/v) P/S, as well as CHIR, PMA and AA. Neuronal progenitor cells remained in glucose-or galactose-based media for 48 hours before proceeding with the Seahorse protocol described above. Either glucose or galactose (always 25mM) were also added to the Seahorse assay medium (XF DMEM pH 7.4) used during the entire procedure.

### Statistical analysis

The predictive accuracy of the PRS model was determined using the area under the receiver operating curve (AUC, pROC R package). A higher AUC indicates a better PRS model that can differentiate between PD cases and controls. We compared the PRS distribution in HC vs iPD subjects using the non-parametric Wilcoxon rank-sum test. To further examine if PRS might predict PD risk, a logistic regression model was used to calculate the odds ratio (OR). Gender, age at assessment and the first three PCs from the population stratification were included as covariates. Individuals were stratified into three groups based on *OXPHOS*-PRS percentiles: low (< 10%), intermediate (10 – 90%) and high (> 90%). Comparison of PD-specific clinical outcomes, AAO and disease duration between individuals in the high and low *OXPHOS*-PRS groups was performed using the Welch’s t-test. Statistical analyses were done in R (v4.0.4) and the *p*-values adjusted using False Discovery Rate (FDR-adj *p* < 0.05) correction (Benjamini-Hochberg method) for the number of independent tests. In functional assays (all involving more than two groups), statistical significance was assessed using one-way ANOVA. For experiments involving two independent variables, two-way ANOVA considering interaction effects has been used. In both cases, Tukey’s post hoc correction for multiple comparisons was applied following significant ANOVA tests. Statistical analyses of functional data were performed using the GraphPad Prism software (v9.4.0). Data with error bars are represented as mean *±* SEM and are representative of at least three independent replicates.

## Results

### Association of common variants in mitochondrial genes with PD risk

To gain essential knowledge on the mitochondria-related “missing heritability” in iPD, we analyzed the distribution of mitoPRSs in two distinct case-control datasets, namely the Luxembourg Parkinson’s Study [15] and COURAGE-PD [16,17] cohorts. After filtering and QC, the final dataset for the Luxembourg Parkinson’s Study comprised 412 iPD patients and 576 HC. iPD patients were older than the HC (67.5±10.9 *vs* 59.1±12.2 years, *p* < 0.01) with a mean AAO of 62.2±11.8 years and a mean disease duration of 5.4±5.0 years (**Table 1**). Using common SNP data from the PD GWAS summary statistics shared by Nalls and colleagues [4], we first calculated the genome-wide PRS for the Luxembourg Parkinson’s Study and found a significant association with PD compared to HC (OR=1.57 per standard deviation; False Discovery Rate adjusted *p*-value, FDR-adj *p* = 8e-09, Area Under the Curve, AUC=0.62, **Fig. 1A**). To capture the cumulative effect of common variants in nuclear-encoded mitochondrial genes on PD risk, we then selected three general mitochondrial gene sets: Human MitoCarta 3.0 [22], and two additional gene lists previously used for mitoPRSs calculation by Billingsley and colleagues [14]. The degree of overlap between the three gene sets is shown in **Supplementary Fig. 1**. Strikingly, all three mitochondrial gene sets were significantly associated with PD (FDR-adj *p* < 0.05, **Fig. 1B**): (i) Billingsley I (OR=1.18[1.02-1.35]), (ii) Billingsley II (OR=1.26[1.09-1.46]) and (iii) MitoCarta 3.0 (OR=1.24[1.08-1.44]). Next, we applied the same approach to distinct mitochondrial pathways, aimed at obtaining PRSs related to specific mitochondrial alterations possibly involved in PD pathogenesis. We, therefore, selected six potentially relevant mitochondrial pathways, containing groups of genes annotated in the corresponding Molecular Signatures Database (MsigDB) sub-collections (**Supplementary Table 1**), and calculated mitochondria-specific PRSs. As shown in **Fig. 1C**, only variants in the *OXPHOS* gene set were significantly associated with a higher PD risk in the Luxembourg Parkinson’s Study (OR=1.31[1.14-1.50], FDR-adj *p* = 5.4e-04). To evaluate genetic risk without considering the potential contribution of variants in known PD loci included in the *OXPHOS* gene list (i.e., *PINK1*, *SNCA* and *PARK7,* as defined by the MDS gene classification list for PD, https://www.mdsgene.org), we ran the PRS analysis for the significant *OXPHOS* pathway after the exclusion of these genes. In this setting, the OR was slightly reduced (OR=1.24[1.08-1.43], FDR-adj *p* = 5.3e-03), but the association with PD risk remained significant (**Fig. 1D**). To take into account SNPs in potential gene regulatory elements, we have also calculated pathway-specific mitoPRSs after extending boundaries upstream (35kb) and downstream (10kb) of each gene [31]. Similar results as before were observed, except for the Billingsley I gene set that was no longer significant (OR=1.15[1.00-1.32], FDR-adj *p* = 0.08). The other mitochondrial gene sets were still significantly associated with PD: Billingsley II (OR=1.21[1.05-1.39], FDR-adj *p* = 1.4e-02), MitoCarta3.0 (OR=1.27[1.10-1.46], FDR-adj *p* = 2.5e-03), as well as *OXPHOS* (OR=1.33[1.16-1.53], FDR-adj p = 2.7e-04) (**Supplementary Fig. 2**). Of note, using common risk alleles associated with other diseases (i.e., Type 2 Diabetes, Schizophrenia, and Alzheimer’s disease GWAS summary statistics) [24–26] for the calculation of mitoPRSs in the imputed Luxembourg Parkinson’s Study cohort, we found that none of them was significantly associated with an increased PD risk (**Supplementary Fig. 3**), thus confirming the specificity of our findings based on PD GWAS common SNPs.

**Figure 1:**
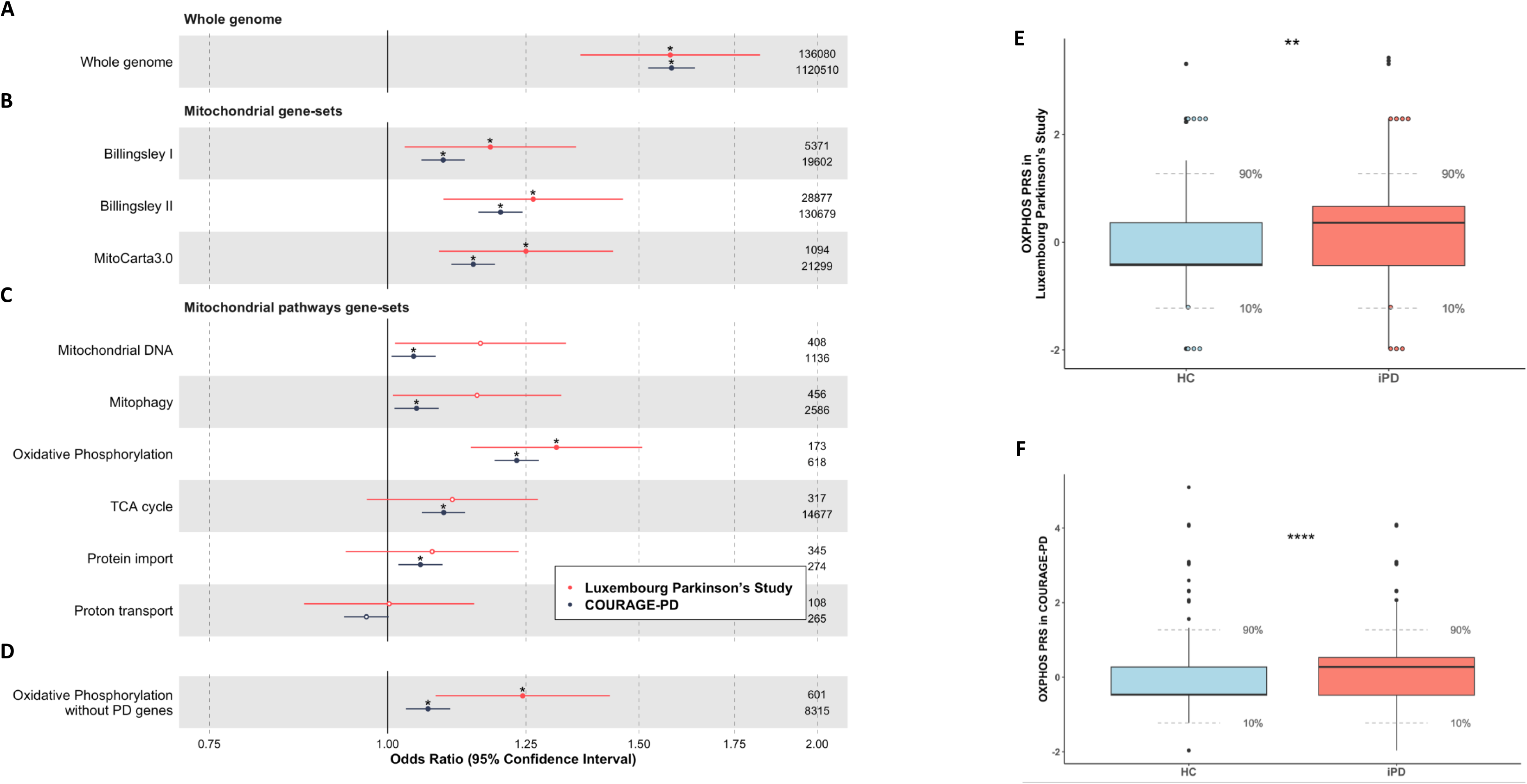
Common variants in mitochondrial genes are associated with higher PD risk. (**A-D**) Forest plots of the odds ratio (OR) and 95% confidence interval for the whole genome (**A**), three different mitochondrial gene sets (**B**), and six selected mitochondrial pathways (**C**) polygenic risk scores (PRSs) regressed with PD diagnosis in the Luxembourg Parkinson’s Study (in red) and COURAGE-PD (in dark blue) cohorts. OR for the significant *OXPHOS* pathway was also plotted upon removal of PD-related loci (i.e., *PINK1*, *SNCA* and *PARK7*) from the *OXPHOS* gene list (**D**). Only significant FDR-adjusted *p*-values are shown on top of each point (* *p* < 0.05). The number of SNPs used to calculate each PRS is shown on the right side. (**E-F**) Distribution of standardized *OXPHOS*-PRS in iPD patients (iPD, red) and healthy controls (HC, light blue) from the Luxembourg Parkinson’s Study (**E**) and COURAGE-PD (**F**) cohorts. The PRS distribution between iPD and HC was significantly different in both datasets (** Wilcoxon test *p*-value < 0.01; **** Wilcoxon test *p*-value < 0.0001). Colored dots in **E** indicate PRS values of iPD cases and HC in the highest and lowest *OXPHOS*-PRS range (10^th^ and 90^th^ percentile, n=4 for each group) whose primary skin fibroblasts were available for functional studies.

Next, we used the 21 cohorts of the COURAGE-PD consortium (**Supplementary Table 2**) [16,17] as a larger replication dataset to validate the results obtained in the exploratory Luxembourg Parkinson’s Study. After filtering and QC, the COURAGE-PD dataset comprised 7270 iPD cases and 6819 HC. Demographic features of COURAGE-PD are reported in **Table 1**. The genome-wide PRS as well as all mitoPRSs tested, including *OXPHOS* (OR=1.23[1.18-1.27], FDR-adj *p* = 1.5e-29), were significantly associated with PD risk in COURAGE-PD (**Fig. 1A-D)**. Again, *OXPHOS*-PRS remained significantly associated with PD risk upon exclusion of the PD-related *PINK1*, *SNCA* and *PARK7* genes from the *OXPHOS* gene list (OR=1.06[1.02-1.10], FDR-adj *p* = 5.6e-04) (**Fig. 1D**).

**Table 1:** Demographic data of iPD patients and healthy controls (HC) from the Luxembourg Parkinson’s Study and COURAGE-PD cohorts.

|  | Luxembourg Parkinson's Study |  | COURAGE-PD |  |
| --- | --- | --- | --- | --- |
|  | iPD | HC | iPD | HC |
| **n** | 412 | 576 | 7270 | 6819 |
| **Age at assessment**<br>(years, mean ± SD) | 67.5±10.9 | 59.1±12.2 | 67.4±11.2 | 66.4±11.8 |
| **Age at onset**<br>(years, mean ± SD) | 62.2±11.8 | - | 58.7±11.6 | - |
| **Disease duration**<br>(years, mean ± SD) | 5.4±5.0 | - | 8.6±6.5 | - |
| **Sex**<br>(% male/female) | 67.9/32.1 | 56.9/43.1 | 60.1/39.9 | 45.3/54.7 |

Means and percentages were calculated after filtering and quality controls of genotyping data.

For both the Luxembourg Parkinson’s study and COURAGE-PD datasets we have also assessed the phenotypic variance and the predictive accuracy of each mitoPRS model, using the R2 metric and the AUC analysis, respectively. Strikingly, among the tested mitochondrial pathways, only the *OXPHOS*-PRS model significantly predicted PD status in both, the Luxembourg Parkinson’s study (R2 = 0.015, FDR-adj *p* = 0.003; AUC = 0.56) and COURAGE-PD (R2 = 0.010, FDR-adj *p* = 3.4e-33; AUC = 0.56) cohorts (**Supplementary Fig. 4** and **Supplementary Table 3**). Finally, to correct for the case-control ratio imbalance, we have also measured the phenotypic variance after adjusting for an estimated PD prevalence of 0.005 [32]. The resulting adjusted R2 (R2*) for the *OXPHOS*-PRS model was still the highest score among the tested pathways in both, the Luxembourg Parkinson’s study (R2* = 0.0058) and COURAGE-PD (R2* = 0.00366) cohorts (**Supplementary Table 3**).

The PRS distribution between iPD and HC was significantly different both in the Luxembourg Parkinson’s Study (Wilcoxon test *p*-value = 1.4e-03, **Fig. 1E**) and in the COURAGE-PD cohorts (Wilcoxon test *p*-value = 2.2e-16, **Fig. 1F**).

To determine whether the *OXPHOS*-PRS could represent a pathophysiology relevant tool to stratify iPD patients based on mitochondrial dysfunction, we sought to functionally validate predicted phenotypes in patient-derived cellular models. Participants were arbitrary grouped into low-, medium-, or high-risk groups according to the PRS distribution in lower and higher deciles [33,34]. Decile cut-offs of 10% and 90%, corresponding to the lowest and highest PD odds ratios, have been chosen for the subsequent functional studies (**Supplementary Fig. 5**). Thus, iPD patients showing the highest or lowest *OXPHOS*-PRSs (10th and 90th percentile, respectively) were identified among the Luxembourg Parkinson’s Study participants and available primary skin fibroblasts from these individuals (n=4 from each group) were used in functional experiments. In parallel, fibroblasts from HC with extreme *OXPHOS*-PRS values (10th and 90th percentile, n=4 for each group) were selected as reference for the comprehensive mitochondrial phenotyping performed in iPD patients’ cells (**Fig. 1E** and **Supplementary Table 4**).

### Mitochondrial oxygen consumption is significantly elevated in primary skin fibroblasts from iPD patients with high *OXPHOS*-PRS

To functionally validate the association between *OXPHOS*-PRS and PD risk in patient-derived cells, we first assessed mitochondrial respiratory chain performance using the Seahorse technology, which provides a comprehensive overview of mitochondrial bioenergetics by measuring oxygen consumption rates (OCRs) under basal conditions and after targeted inhibition of specific respiratory chain complexes [29]. Strikingly, basal respiration, proton leak and ATP-linked respiration were all significantly enhanced in fibroblasts from iPD patients with high *OXPHOS*-PRS compared to low *OXPHOS*-PRS cells, reaching OCR levels well beyond the reference threshold defined by fibroblasts of healthy individuals (**Fig. 2** and **Supplementary Fig. 6**). In this setting, maximal respiration was not significantly different between high and low *OXPHOS*-PRS iPD fibroblasts, indicating a reduced spare respiratory capacity in the high *OXPHOS*-PRS iPD group. Of note, all respiratory parameters analyzed did not significantly vary between HC with high or low *OXPHOS*-PRS (**Fig. 2** and **Supplementary Fig. 6**). Likewise, biogenesis of respiratory chain complexes (RCC) did not differ significantly between high and low *OXPHOS*-PRS groups – both in HC and in iPD patients – as revealed by immunoblot analysis of RCC protein subunits (**Supplementary Fig. 7**). Collectively, these findings suggest that fibroblasts of iPD patients with high *OXPHOS*-PRS display features of mitochondrial hyperactivity already under steady-state conditions, a phenotype that appears to be independent of RCC protein expression.

**Figure 2:**
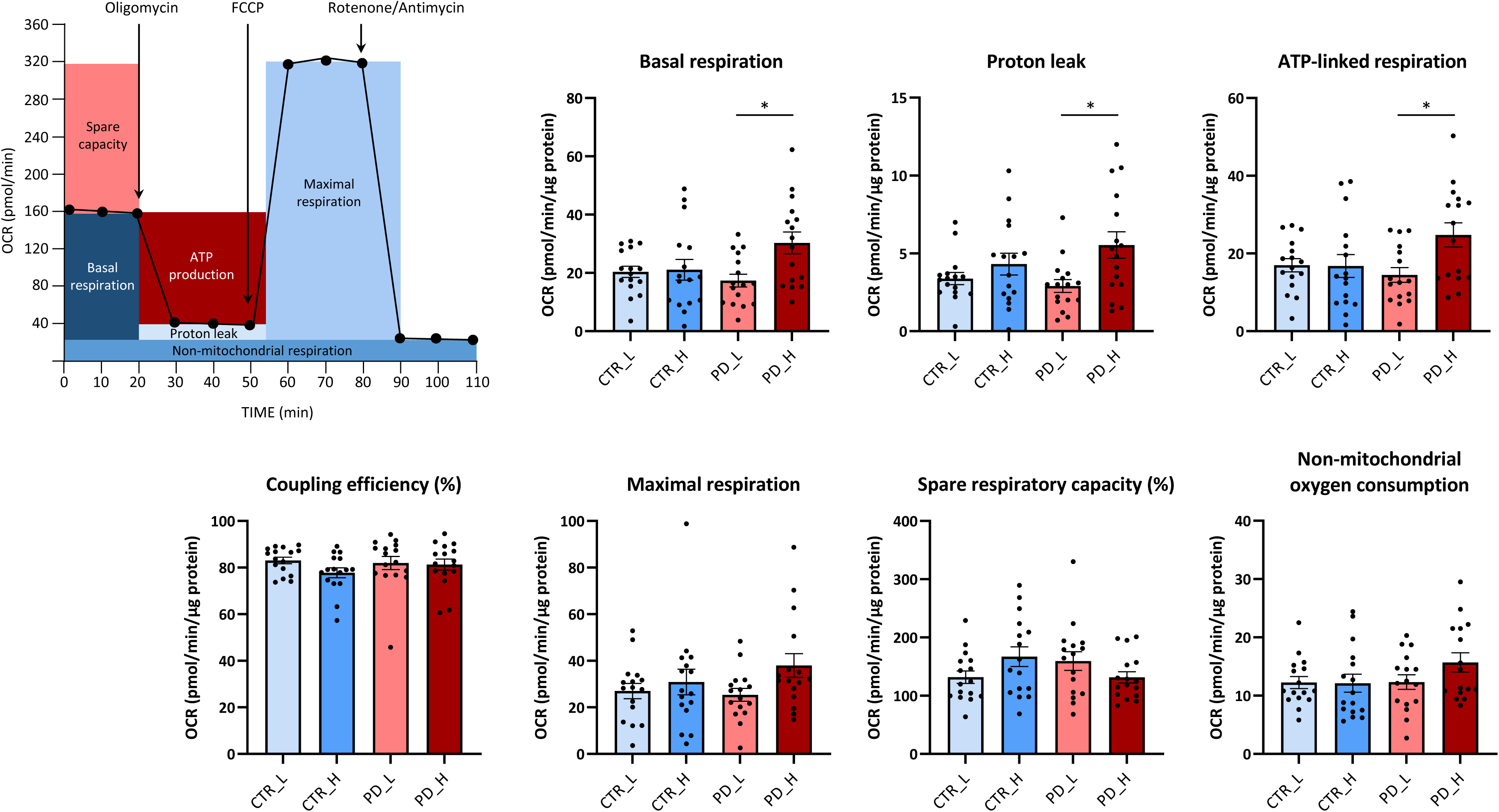
Analysis of mitochondrial respiration in primary skin fibroblasts from iPD patients and HC stratified based on *OXPHOS*-PRS. Oxygen consumption rates (OCRs) were measured under basal conditions and after targeted inhibition of specific respiratory chain complexes by using a standard Seahorse Mito Stress test. Histobars represent the pooled means ± SEM of four independent experiments performed in four distinct fibroblast lines established from Luxembourg Parkinson’s Study participants (healthy controls vs iPD patients) with high (CTR_H; PD_H) or low (CTR_L; PD_L) *OXPHOS*-PRS. Six technical replicates per group (distinct Seahorse wells) were performed in each experiment. *P < 0.05. One-way ANOVA correcting for multiple comparisons using the Tukey’s post hoc test.

### Other functional readouts of mitochondrial activity are not significantly altered in fibroblasts from iPD patients stratified based on *OXPHOS*-PRS

To assess whether changes in mitochondrial respiration were accompanied by differences in reactive oxygen species (ROS) production, fibroblasts from both iPD patients and HC stratified according to their individual *OXPHOS*-PRS were subjected to CellROX staining. Surprisingly, increased ROS levels were detected in HC from the high *OXPHOS*-PRS group compared to low-risk individuals, but not in iPD patients, arguing against a causal link between elevated OXPHOS and ROS accumulation in these cells (**Fig. 3A** and **Supplementary Fig. 8A**). Given the crucial role of the electrochemical gradient across the inner mitochondrial membrane in generating the proton motive force used by the OXPHOS machinery to synthesize ATP [35], we also measured mitochondrial membrane potential (ΔΨ_m_) in primary skin fibroblasts from each *OXPHOS*-PRS group. To this end, cell lines were stained with Tetramethylrhodamine ethyl ester (TMRE), a fluorescent dye that specifically accumulates in active, polarized mitochondria, followed by high-throughput automated confocal microscopy analysis. Treatment with the OXPHOS uncoupler carbonyl cyanide 3-chlorophenylhydrazone (CCCP), known to induce mitochondrial depolarization, was used as a positive control for decreased ΔΨ_m_. As expected, CCCP-treated cells all failed to accumulate the dye within mitochondria, and therefore displayed a robust reduction of TMRE fluorescence. However, no significant differences in ΔΨ_m_ were observed between untreated fibroblasts from both HC and iPD patients previously stratified according to their low or high *OXPHOS*-PRS (**Fig. 3B** and **Supplementary Fig. 8B**). OXPHOS can also be affected by abnormal mitochondrial fission/fusion processes, as revealed by changes in ETC activity and respiratory function in response to dynamic transitions of organelle morphology – from fragmented to elongated, and vice versa [36,37]. Again, we observed no difference in morphological parameters such as mitochondrial *aspect ratio* and *form factor* in fibroblasts derived from HC and iPD patients with high or low *OXPHOS*-PRS (**Fig. 3C** and **Supplementary Fig. 8C)**. mtDNA copy number, transcription/replication rates and deletions, which could impinge on ETC activity by altering the expression and stoichiometric assembly of mtDNA-encoded RCC subunits [12,38,39], were also not significantly different between high and low *OXPHOS*-PRS groups (**Supplementary Fig. 9**). Finally, based on the emerging link between mitochondrial dysfunction and inflammation in PD [40,41], we measured interleukin-6 (IL-6) levels in blood plasma samples from the Luxembourg Parkinson’s Study participants (both HC and iPD patients) with the highest or lowest *OXPHOS*-PRS. Indeed, we recently found increased IL-6 levels in the serum of PD patients carrying biallelic mutations in *PINK1* or *PRKN* [40], a phenotype mainly associated with impaired mitophagy and dependent on activation of the pro-inflammatory cGAS-STING pathway [41]. Here, we did not observe significant differences in plasma IL-6 levels between high and low *OXPHOS*-PRS groups (**Fig. 3D**).

**Figure 3:**
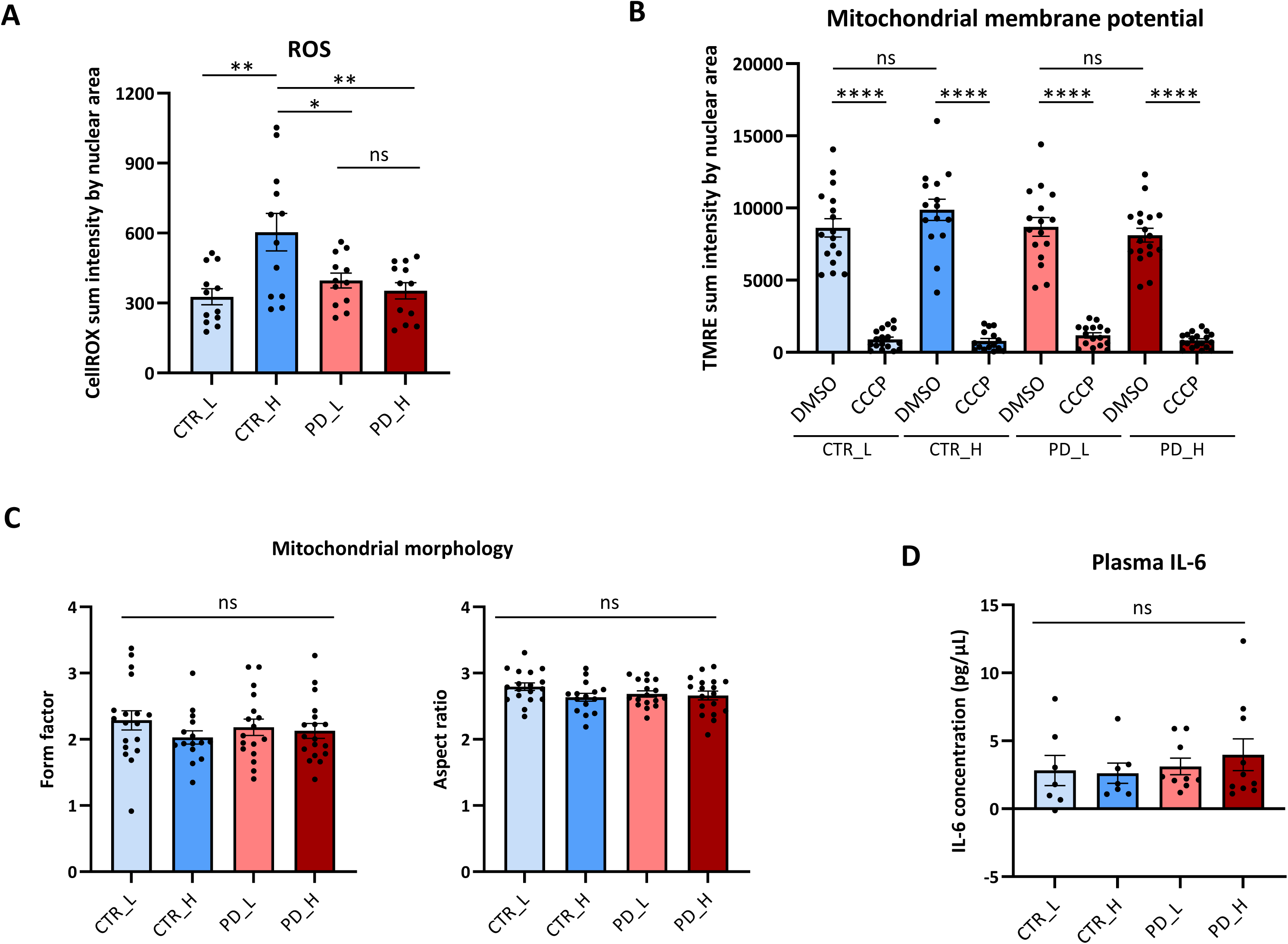
Assessment of additional readouts of mitochondrial activity in primary skin fibroblasts and plasma samples from iPD patients and HC stratified based on *OXPHOS*-PRS. (**A-C**) High-throughput confocal microscopy analyses in primary skin fibroblasts derived from Luxembourg Parkinson’s Study participants with high (CTR_H; PD_H) or low (CTR_L; PD_L) *OXPHOS*-PRS. (**A**) ROS levels were quantified by normalizing the CellRox mean fluorescence intensity against the nuclear area, as defined by the Hoechst staining. Histobars represent the pooled means ± SEM of three independent experiments performed in four distinct fibroblast lines for each group. (**B**) Mitochondrial membrane potential (ΔΨ_m_) was measured after normalization of TMRE mean fluorescence intensity by the nuclear area. Treatment with the OXPHOS uncoupler carbonyl cyanide 3-chlorophenylhydrazone (CCCP), known to induce mitochondrial depolarization, was used as positive control for decreased ΔΨ_m_. Dimethyl sulfoxide (DMSO) was used as a vehicle. Histobars represent the pooled means ± SEM of at least three independent experiments performed in four distinct fibroblast lines for each group. (**C**) Morphometric analysis of the mitochondrial network. *Form factor* and *aspect ratio* were quantified as described previously [65]. Histobars represent the pooled means ± SEM of at least three independent experiments performed in four distinct fibroblast lines for each group. (**D**) Measurement of IL-6 levels in plasma samples obtained from Luxembourg Parkinson’s Study participants (healthy controls vs iPD patients) with high (CTR_H, n=7; PD_H, n=10) or low (CTR_L, n=7; PD_L, n=9) *OXPHOS*-PRS. ** *p* < 0.01. ns = not significant.

### Functional validation of *OXPHOS-PRS* in iPSC-derived neuronal progenitor cells

To validate our *OXPHOS*-PRS approach in more disease-relevant patient-derived models, we reprogrammed fibroblast lines into induced pluripotent stem cells (iPSCs). Given that fibroblasts from HC did not display major differences when comparing low-and high-risk groups, we decided to focus on iPD patients only and successfully reprogrammed two lines per group into iPSCs (#17162 and #18250 for the low *OXPHOS*-PRS; #11043 and #15092 for the high *OXPHOS*-PRS). The four iPSC lines all displayed no chromosomal aberrations, genetic background identical to the original skin fibroblasts, and typical stem cell features (**Supplementary Fig. 10A-B**). Different from fibroblasts, iPSCs all expressed high RNA levels of the stemness markers *OCT3/4* and *NANOG* (**Supplementary Fig. 10C**). Pluripotency was further corroborated by a robust nuclear staining for the Nanog protein (**Supplementary Fig. 10D**). Next, newly generated iPSC lines were differentiated into smNPCs [27], and then subjected to an assessment of mitochondrial respiration. Seahorse-based bioenergetics analyses in smNPCs did not reveal major differences in OCRs between the low and high *OXPHOS*-PRS groups (**Fig. 4**), likely due to their high glycolytic activity and low reliance on OXPHOS metabolism [42]. In accordance with this hypothesis, shifting smNPCs from glucose to galactose medium as carbon source dramatically increased basal and ATP-linked respiration, indicative of a metabolic switch from glycolysis to OXPHOS (**Fig. 4**). In this setting, smNPCs derived from iPD patients with high *OXPHOS*-PRSs displayed a significant decrease of basal and ATP-linked respiration compared to the low *OXPHOS*-PRSs group, comparable to that observed in smNPCs from patients with autosomal recessive PD caused by mutations in the *PINK1* gene (**Fig. 4**).

**Figure 4:**
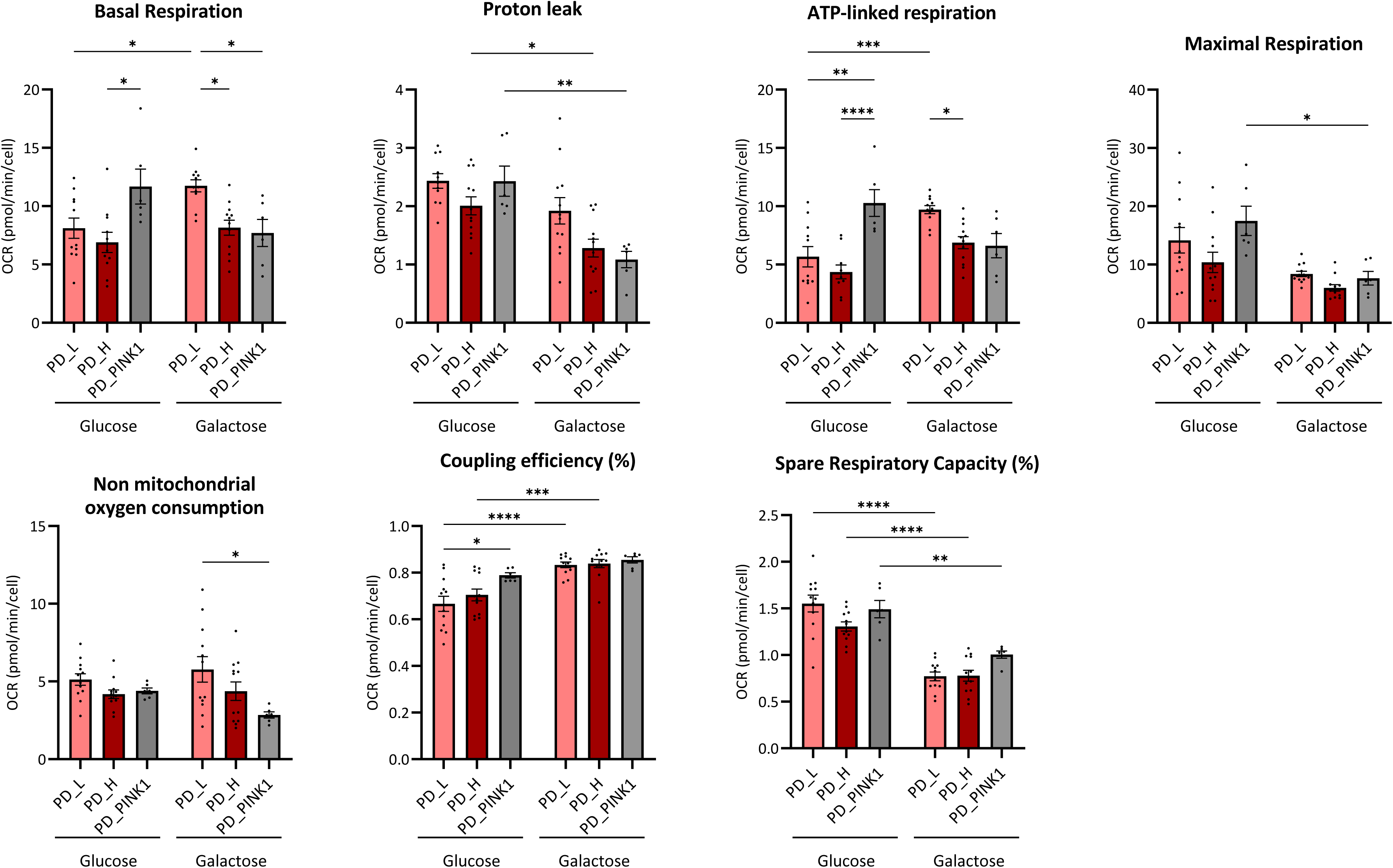
Functional validation of *OXPHOS-PRS* in iPSC-derived neuronal progenitor cells. Oxygen consumption rates (OCRs) were measured under basal conditions and after targeted inhibition of specific respiratory chain complexes by using a standard Seahorse Mito Stress test. Histobars represent the pooled means ± SEM of either 6 (PD_L and PD_H) or 3 (PD_PINK1) independent experiments performed in two distinct smNPC lines per group, cultivated either in glucose or galactose medium. Eight technical replicates per group (distinct Seahorse wells) were performed in each experiment. *P < 0.05; **P < 0.01; ***P < 0.001; ****P < 0.0001. Two-way ANOVA correcting for multiple comparisons using the Tukey’s post hoc test.

### Association of *OXPHOS*-PRSs with PD-specific clinical outcomes

Finally, we sought to investigate whether our functionally validated *OXPHOS*-PRS approach could be used to define distinct phenotypic patterns among iPD patients. To this end, we first analyzed eight PD-specific motor and non-motor clinical scores in iPD patients from the Luxembourg Parkinson’s Study stratified according to their *OXPHOS*-PRS, namely the Movement Disorder Society update of the Unified Parkinson’s Disease Rating Scale I (MDS-UPDRS I), MDS-UPDRS II, MDS-UPDRS III, MDS-UPDRS-IV, the Quality of Life questionnaire (PDQ39), the L-dopa-equivalent daily dose (LEDD), the Scales for Outcomes in Parkinson’s Disease – Autonomic Dysfunction (SCOPA-AUT) and the Montreal Cognitive Assessment (MoCA). The whole-genome PRS was used as reference. In general, the trend of all clinical outcomes analyzed was similar to that observed for the whole-genome PRS, and none of them was significantly different between iPD patients with high or low *OXPHOS*-PRS (**Supplementary Fig. 11**). However, iPD patients with high *OXPHOS*-PRS displayed an earlier AAO compared to low-risk patients, a phenotype particularly evident in the larger COURAGE-PD dataset (FDR-adj *p* = 0.015, **Fig. 5**).

**Figure 5:**
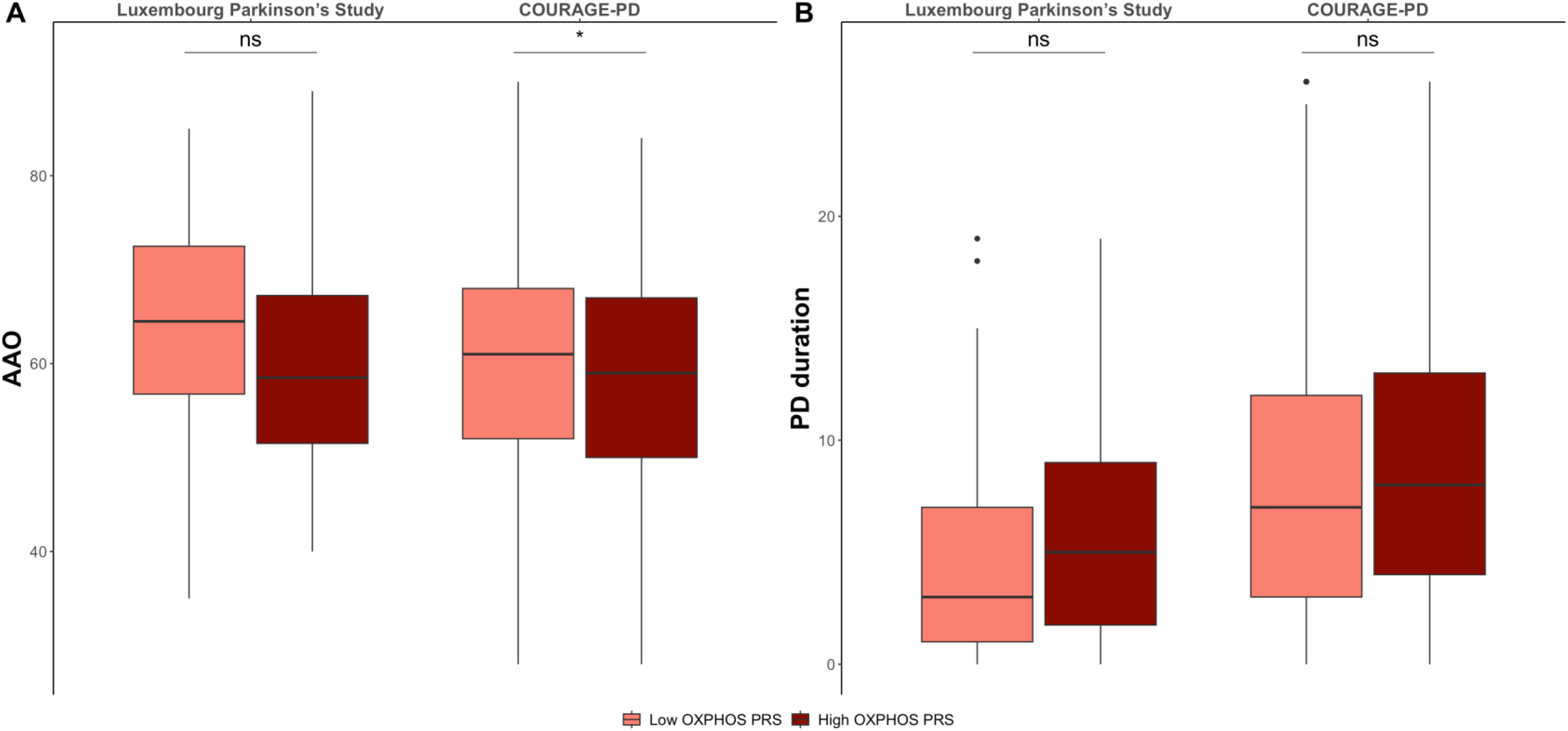
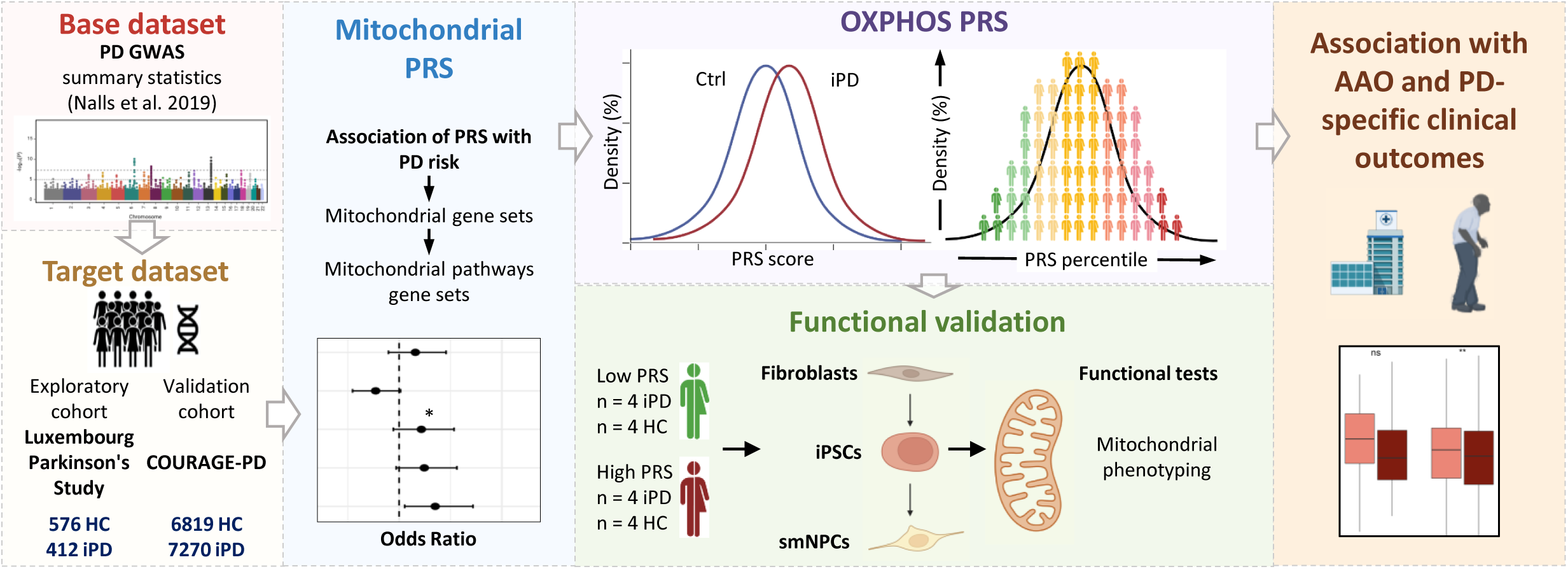
Association of *OXPHOS*-PRSs with age at PD onset and disease duration. Comparison of mean age at PD onset (AAO, **A**) and mean disease duration (**B**) in iPD patients from the Luxembourg Parkinson’s Study and COURAGE-PD cohorts with high or low *OXPHOS*-PRS. * *p* < 0.05, ns = not significant.

## Discussion

Despite recent progress in the generation and analysis of large-scale genotyping data, deciphering the complex genetic architecture of PD is still a major challenge. Low-penetrance genetic variants that significantly increase the risk to develop PD have been identified in sporadic cases (e.g., *GBA* L444P and *LRRK2* G2019S), but typical hallmarks of PD heritability can also be observed in families with unknown genetic causes [43], which suggests a potential involvement of genetic factors in up to 30% of identified PD cases [44].

Missing heritability in PD and the clear association between mitochondrial dyshomeostasis and neurodegeneration led us to investigate the role of disease-associated SNPs in genes regulating mitochondrial function. Thus, we performed association analyses targeting common variants in nuclear-encoded mitochondrial genes and analyzed the mitoPRS distribution in two distinct iPD case-control studies, namely the Luxembourg Parkinson’s Study and the COURAGE-PD consortium. After testing different general mitochondrial gene sets, we found that mitoPRSs were always significantly associated with increased PD risk in both cohorts, thus confirming the results obtained by other groups [14,45]. Of note, PRSs can also be applied to subsets of genes controlling distinct, disease-related, cellular activities (e.g., mitochondrial quality control or lysosomal function in PD), aimed at identifying specific pathways implicated in the pathological phenotype [32]. Using this approach, Paliwal and colleagues recently demonstrated that certain mitochondrial pathways – including *OXPHOS* among others – were significantly associated with increased Alzheimer’s disease risk [46]. In a similar manner, here, we customized gene lists for six mitochondrial pathways relevant for PD and calculated pathway-specific mitoPRSs. Strikingly, only common variants in genes regulating *OXPHOS* were significantly associated with increased PD risk in the Luxembourg Parkinson’s Study and COURAGE-PD cohorts, reinforcing the notion of impaired mitochondrial respiration as a crucial aspect in PD pathogenesis.

PRS-based approaches appear increasingly useful to dissect genetically heterogeneous disorders like PD and to identify pathophysiology-relevant iPD subtypes [5,47–49]. However, the major unmet need in the field is the functional validation of PRSs in patient-based models, which would legitimate the effectiveness of genetically predicted cellular phenotypes thus paving the way for precision medicine therapeutic approaches [50]. Here, we functionally validated individual *OXPHOS*-PRS profiles in both primary skin fibroblasts and the corresponding iPSC-derived neuronal progenitor cells established from genetically stratified iPD patients, detecting significant differences in mitochondrial respiration between high-and low-risk groups. In primary skin fibroblasts, OCRs were significantly elevated in the high *OXPHOS*-PRS group – especially basal and ATP-linked respiration. It has been postulated that a similar phenotype, indicative of steady-state mitochondrial hyperactivity, may be an early event in PD pathogenesis, as suggested by increased neuronal viability and improved motor function in worms treated with the mitochondrial complex I inhibitor metformin [51,52]. Based on this model, hyperactive mitochondria may not only reduce the ability of the *OXPHOS* machinery to cope with a sudden increase in energy demand (e.g., in response to acute cellular stress), but also cause an excessive accumulation of ROS, which in turn increases oxidative damage and ultimately induces neurodegeneration. In this scenario, reduced ETC complex activity and impaired mitochondrial respiration – usually observed in peripheral tissues and *post-mortem* brains from PD patients – would be late endpoint phenotypes, caused by ROS-mediated damage of mitochondrial complex I [53]. However, in the current study, increased mitochondrial respiration in fibroblasts with high *OXPHOS*-PRS was not accompanied by enhanced ROS production, likely due to the activation of compensatory mechanisms resulting in increased scavenging capacity [54]. High OCRs may also reflect the specific metabolism of the cells or tissues studied, including their ability to cope with mitochondrial dysfunction and to ensure ATP production. Similar to our findings, mitochondrial respiration was found to be significantly higher in Parkin-mutant fibroblasts compared to controls [55,56], and mitochondrial hyperactivity was also observed in lymphoblasts from iPD patients [57]. In these studies, high OCRs were not accompanied by alterations in mitochondrial membrane potential or mtDNA copy number [55,57], which is in line with our observations and further reinforces our stratification approach based on nuclear-encoded mitochondrial genes.

Results obtained in dermal fibroblasts are certainly informative and help to identify biological processes and molecular alterations also playing a relevant role in the neuronal environment, but their gene expression profile, metabolic adaptation to stress and signaling pathways strongly differ from neurons [58]. Moreover, the epigenetic changes that occur during somatic cell reprogramming to iPSCs and the subsequent differentiation into neuronal precursors could lead to opposing results between primary skin fibroblasts and smNPCs [59]. In accordance with this, mitochondrial respiration in iPSC-derived neuronal progenitor cells established from the corresponding reprogrammed fibroblasts was very similar between low and high *OXPHOS*-PRS groups. However, when forced to use OXPHOS instead of glycolysis to produce ATP (i.e., in galactose medium), smNPCs derived from iPD patients with high *OXPHOS*-PRSs displayed a significant reduction in their OCRs compared to the low *OXPHOS*-PRS group. Importantly, degree of mitochondrial dysfunction in smNPCs from iPD patients with high *OXPHOS*-PRSs is comparable to that observed in cells from autosomal recessive PD patients carrying *PINK1* loss-of-function mutations (see **Fig. 4**). Of note, *PINK1*-PD is typically characterized by early onset and long disease duration, a phenotype that – according to our findings – is also observed in iPD patients with high *OXPHOS*-PRSs. This further corroborates the effectiveness of our functionally validated genetic stratification approach and highlights the possibility to include iPD patients with high OXPHOS-PRS in future clinical trials with mitochondrially active drugs.

Based on insights into the pathophysiology of neurodegeneration in PD, mitochondria early emerged as a potential therapeutic target for disease-modifying treatments (DMT) [60]. However, all clinical trials focusing on mitochondrially active compounds in PD (e.g., coenzyme Q10) failed, raising doubts as to the validity of the concept of applying DMT to unselected patient groups given the complex and heterogeneous nature of PD [61]. Therefore, more recently, first clinical trials have been initiated, which apply genetic stratification strategies to define subgroups of PD patients, i.e., carriers of rare mutations in genes linked to mitochondrial dyshomeostasis and early-onset forms of PD [62]. If such approaches were successful, it would be still a very limited subgroup of PD patients that could benefit from these efforts. Thus, our functionally validated genetic stratification approach may address current needs for more focused DMT trials directed to a broader group of patients with an underlying, genetically defined, mitochondrial burden. Such an approach, coupled with compounds that showed ample preclinical and clinical evidence of targeting mitochondrial OXPHOS defects [63], may even be effective if a given drug previously failed in trials including unselected PD patient groups. Finally, since increased mitochondrial genetic risk is often associated with early-onset PD forms, individuals with very high *OXPHOS*-PRS could also represent a suitable target for preventative therapies.

Although the association between *OXPHOS*-PRS and PD risk is weaker than that observed for the whole-genome PRS (see **Fig. 1**), narrowing specific pathways – with fewer genes, but still significant effect – can shed new light on potential molecular mechanisms underlying PD pathogenesis. Thus, different from the whole-genome PRS, we believe that our genetic stratification approach based on *OXPHOS*-PRS represents a “precision tool” that may have a considerable impact on personalized medicine in PD. Despite this, we have to acknowledge that our study has some limitations. The relatively small size of the Luxembourg Parkinson’s Study cohort decreased statistical power of our analyses, therefore one could speculate that individuals with extreme *OXPHOS*-PRS values may not be in the top/bottom ranks of a larger cohort. This would explain why the inverse association between *OXPHOS*-PRS and AAO was statistically significant in iPD patients from the large COURAGE-PD dataset, whereas differences, albeit consistent, did not reach statistical relevance in the Luxembourg Parkinson’s Study. On the other hand, available genetic information, clinical data and, at least in part, biospecimens from all the subjects followed longitudinally in the Luxembourg Parkinson’s Study, make this monocentric cohort quite unique, and the concomitant assessment of PD phenotypes, environmental risk factors and omics data could be integrated with *OXPHOS*-PRS to improve iPD patients’ stratification accuracy. Follow-up studies on additional large and well-phenotyped PD cohorts are warranted to further explore the link between genetic stratification using *OXPHOS*-PRSs and the development of PD-associated clinical outcomes. Another possible limitation of our study is that skin biopsies from individuals with the very highest *OXPHOS*-PRSs (three iPD patients and one HC, see **Fig. 1E**) were not available at the time we started the functional experiments, therefore differences in mitochondrial respiration between iPD patients with low and high *OXPHOS*-PRSs, although already significant, might be even more pronounced if we could have included fibroblasts and iPSC-derived neuronal progenitor cells from these individuals. Functional validation of *OXPHOS*-PRSs in best ranked and in a broader number of iPD patients from larger cohorts will be a prerequisite for translating genetic prediction into clinical practice.

## Conclusions

To date, only three studies used PRS-based approaches to assess mitochondrial risk in PD [14,32,45]. Two of them demonstrated that combination of small effect-size common variants in nuclear-encoded mitochondrial genes were significantly associated with higher PD risk [14,45], while the other one failed to confirm this association but showed functional enrichment of mitochondria-related pathways – including OXPHOS [32]. One of these studies also demonstrated an association between mitoPRSs and clinical outcomes [45]. Nevertheless, all these studies did not consider the need to functionally validate mitoPRSs in patient-based cellular models, which is an essential step for translating genetic prediction into clinical practice [50].

In addition, there have been attempts to stratify iPD patients exclusively based on mitochondrial phenotyping assays [64]. However, this type of stratification is time-consuming, labor-intensive, and error-prone; in fact, since most functional assays rely on relative measures, the definition of cut-offs between “mitochondrial” and “non-mitochondrial” PD subtypes is extremely challenging. In this light, the translation of a purely mitochondrial phenotyping approach into clinical trials for the stratification of a large number of patients seems impractical.

Our study follows a different idea: here, the patient stratification is entirely based on genetics and therefore defined by ‘absolute’ measures. This makes our approach robust, cost-effective and high-throughput compatible, as genetics can be easily assessed and provides ‘definitive’ data. Our findings, obtained in two distinct case-control datasets, clearly suggest that only variants in genes regulating oxidative phosphorylation (as defined by the *OXPHOS*-PRS) are significantly associated with an increased risk of developing PD. Importantly, for the first time in the PD field, we functionally validated genetic prediction (i.e., altered oxygen consumption rates) in cellular models derived from the corresponding PD patients. Moreover, and further corroborating our hypothesis, we demonstrated a significant association between *OXPHOS*-PRS and the clinical phenotype similar to what can be expected from known monogenic forms of PD related to mitochondrial defects (e.g., *PINK1*), as defined by an earlier age at PD onset in the high-risk group. Combining genetic analyses, experimental validation studies and clinical assessment, all from the same individuals, certainly goes far beyond existing work, and provides a novel concept for precision medicine that is relevant for physicians and researchers engaged in translating fundamental research into therapeutic intervention for the treatment of neurodegenerative diseases. We are confident that our robust and high-throughput compatible *OXPHOS*-PRS approach may address current needs for more personalized clinical trials directed to iPD patients with an underlying, genetically defined, mitochondrial phenotype, who may benefit from drugs specifically targeting the respiratory chain activity.

## Declarations

### Ethics approval and consent to participate

All participants signed a written informed consent according to the Declaration of Helsinki. Ethical approval for the usage of patient-derived cell lines was given by the National Committee for Ethics in Research, Luxembourg (Comité National d’Ethique dans la Recherche; CNER #201411/05).

### Consent for publication

Not applicable

### Availability of data and materials

Genetic and clinical data for this manuscript are not publicly available as they are linked to the internal regulations of the Luxembourg Parkinson’s Study and COURAGE-PD consortia. Requests for accessing the datasets can be directed to, where dedicated data access committees review the requests and conclude data sharing agreements with prospective users.

## Funding

Luxembourg National Research Fund (C17/BM/11676395 to RK, AG, PM, EG and GA)

Luxembourg National Research Fund (FNR/NCER13/BM/11264123 to RK)

Luxembourg National Research Fund (FNR/P13/6682797 to RK)

Luxembourg National Research Fund (C21/BM/15850547 to GA)

Luxembourg National Research Fund (FNR9631103 to AG)

Luxembourg National Research Fund and German Research Council (FNR/DFG 11250962 to PM and AG)

Luxembourg National Research Fund and German Research Council (FNR/DFG 11676395 to MS and AAKS)

Luxembourg National Research Fund (EU Horizon 2020 programme, grant no. ERAPERMED 2020-314, DIGI-PD to EG).

### Authors’ contribution

Conceptualization: GA, DG, MS, RK, PM, AG

Methodology: GA, ZL, PA, IB, DRB, AAKS

Investigation: GA, ZL, AV, SD, LP

Visualization: GA, ZL, LP

Funding acquisition: GA, EG, MS, RK, PM, AG

Project administration: GA, RK, PM, AG

Supervision: GA, RK, PM, AG, MS

Writing – original draft: GA, ZL

Writing – review & editing: SD, PA, IB, LP, MS, EG, RK, PM, AG

## Supporting information

Supplementary informations

## List of abbreviations

ΔΨ_m_: mitochondrial membrane potential
AA: Ascorbic Acid
AAO: Age at onset
AUC: area under the receiver operating curve
CCCP: carbonyl cyanide 3-chlorophenylhydrazone
CHIR: CHIR 99021
COURAGE-PD: Comprehensive Unbiased Risk Factor Assessment for Genetics and Environment in Parkinson’s Disease
DAN: dopaminergic neuron
DMEM: Dulbecco’s Modified Eagle Medium
DMSO: Dimethyl sulfoxide
DMT: disease-modifying treatments
ETC: electron transport chain
FCCP: carbonyl cyanide p-trifluoromethoxyphenylhydrazone
FDR: False Discovery Rate
GWAS: genome-wide association studies
HC: healthy controls
HI-FBS: heat-inactivated Fetal Bovine Serum
IL-6: interleukin-6
iPD: idiopathic PD
IPDGC: International Parkinson’s Disease Genomics Consortium
iPSCs: induced pluripotent stem cells
LD: linkage disequilibrium
LEDD: L-dopa-equivalent daily dose
MDS-UPDRS: Movement Disorder Society update of the Unified Parkinson’s Disease Rating Scale
mitoPRS: mitochondria-specific polygenic risk score
MoCA: Montreal Cognitive Assessment (MoCA)
MsigDB: Molecular Signatures Database
mtDNA: mitochondrial DNA
NCER-PD: National Centre for Excellence in Research in Parkinson’s Disease
OCRs: Oxygen consumption rates
OR: odds ratio
OXPHOS: oxidative phosphorylation
PC: Principal components
PD: Parkinson’s disease
PDQ39: Parkinson’s Disease Questionnaire
PMA: purmorphamine
PRS: polygenic risk score
P/S: Penicillin-Streptomycin
QC: Quality control
RCC: respiratory chain complexes
ROS: reactive oxygen species
SCOPA-AUT: Scales for Outcomes in Parkinson’s Disease – Autonomic Dysfunction
smNPCs: neuronal progenitor cells
SNP: single nucleotide polymorphisms
TMRE: Tetramethylrhodamine ethyl ester

## Acknowledgements

We would like to thank all participants in the Luxembourg Parkinson’s Study for their important support of our research. Furthermore, we acknowledge the joint effort of the NCER-PD consortium members from the partner institutions Luxembourg Centre for Systems Biomedicine, Luxembourg Institute of Health, Centre Hospitalier de Luxembourg and Laboratoire National de Santé, as well as all members of the international COURAGE-PD consortium.

## NCER-PD consortium

Alexander HUNDT 2, Alexandre BISDORFF 5, Amir SHARIFY 2, Anne GRÜNEWALD 1, Anne-Marie HANFF 2, Armin RAUSCHENBERGER 1, Beatrice NICOLAI 3, Brit MOLLENHAUER 12, Camille BELLORA 2, Carlos MORENO 1, Chouaib MEDIOUNI 2, Christophe TREFOIS 1, Claire PAULY 1,3, Clare MACKAY 10, Clarissa GOMES 1, Daniela BERG 11, Daniela ESTEVES 2, Deborah MCINTYRE 2, Dheeraj REDDY BOBBILI 1, Eduardo ROSALES 2, Ekaterina SOBOLEVA 1, Elisa GÓMEZ DE LOPE 1, Elodie THIRY 3, Enrico GLAAB 1, Estelle HENRY 2, Estelle SANDT 2, Evi WOLLSCHEID-LENGELING 1, Francoise MEISCH 1, Friedrich MÜHLSCHLEGEL 4, Gaël HAMMOT 2, Geeta ACHARYA 2, Gelani ZELIMKHANOV 3, Gessica CONTESOTTO 2, Giuseppe ARENA 1, Gloria AGUAYO 2, Guilherme MARQUES 2, Guy BERCHEM 3, Guy FAGHERAZZI 2, Hermann THIEN 2, Ibrahim BOUSSAAD 1, Inga LIEPELT 11, Isabel ROSETY 1, Jacek JAROSLAW LEBIODA 1, Jean-Edouard SCHWEITZER 1, Jean-Paul NICOLAY 19, Jean-Yves FERRAND 2, Jens SCHWAMBORN 1, Jérôme GRAAS 2, Jessica CALMES 2, Jochen KLUCKEN 1,2,3, Johanna TROUET 2, Kate SOKOLOWSKA 2, Kathrin BROCKMANN 11, Katrin MARCUS 13, Katy BEAUMONT 2, Kirsten RUMP 1, Laura LONGHINO 3, Laure PAULY 1, Liliana VILAS BOAS 3, Linda HANSEN 1,3, Lorieza CASTILLO 2, Lukas PAVELKA 1,3, Magali PERQUIN 2, Maharshi VYAS 1, Manon GANTENBEIN 2, Marek OSTASZEWSKI 1, Margaux SCHMITT 2, Mariella GRAZIANO 17, Marijus GIRAITIS 2,3, Maura MINELLI 2, Maxime HANSEN 1,3, Mesele VALENTI 2, Michael HENEKA 1, Michael HEYMANN 2, Michel MITTELBRONN 1,4, Michel VAILLANT 2, Michele BASSIS 1, Michele HU 8, Muhammad ALI 1, Myriam ALEXANDRE 2, Myriam MENSTER 2, Nadine JACOBY 18, Nico DIEDERICH 3, Olena TSURKALENKO 2, Olivier TERWINDT 1,3, Patricia MARTINS CONDE 1, Patrick MAY 1, Paul WILMES 1, Paula Cristina LUPU 2, Pauline LAMBERT 2, Piotr GAWRON 1, Quentin KLOPFENSTEIN 1, Rajesh RAWAL 1, Rebecca TING JIIN LOO 1, Regina BECKER 1, Reinhard SCHNEIDER 1, Rejko KRÜGER 1,2,3, Rene DONDELINGER 5, Richard WADE-MARTINS 9, Robert LISZKA 14, Romain NATI 3, Rosalina RAMOS LIMA 2, Roseline LENTZ 7, Rudi BALLING 1, Sabine SCHMITZ 1, Sarah NICKELS 1, Sascha HERZINGER 1, Sinthuja PACHCHEK 1, Soumyabrata GHOSH 1, Stefano SAPIENZA 1, Sylvia HERBRINK 6, Tainá MARQUES 1, Thomas GASSER 11, Ulf NEHRBASS 2, Valentin GROUES 1, Venkata SATAGOPAM 1, Victoria LORENTZ 2, Walter MAETZLER 15, Wei GU 1, Wim AMMERLANN 2, Yohan JAROZ 1, Zied LANDOULSI 1.

1 Luxembourg Centre for Systems Biomedicine, University of Luxembourg, Esch-sur-Alzette, Luxembourg
2 Luxembourg Institute of Health, Strassen, Luxembourg
3 Centre Hospitalier de Luxembourg, Strassen, Luxembourg
4 Laboratoire National de Santé, Dudelange, Luxembourg
5 Centre Hospitalier Emile Mayrisch, Esch-sur-Alzette, Luxembourg
6 Centre Hospitalier du Nord, Ettelbrück, Luxembourg
7 Parkinson Luxembourg Association, Leudelange, Luxembourg
8 Oxford Parkinson’s Disease Centre, Nuffield Department of Clinical Neurosciences, University of Oxford, Oxford, UK
9 Oxford Parkinson’s Disease Centre, Department of Physiology, Anatomy and Genetics, University of Oxford, Oxford, UK
10 Oxford Centre for Human Brain Activity, Wellcome Centre for Integrative Neuroimaging, Department of Psychiatry, University of Oxford, Oxford, UK
11 Center of Neurology and Hertie Institute for Clinical Brain Research, Department of Neurodegenerative Diseases, University Hospital Tübingen, Tübingen, Germany
12 Paracelsus-Elena-Klinik, Kassel, Germany
13 Ruhr-University of Bochum, Bochum, Germany
14 Westpfalz-Klinikum GmbH, Kaiserslautern, Germany
15 Department of Neurology, University Medical Center Schleswig-Holstein, Kiel, Germany
16 Department of Neurology Philipps, University Marburg, Marburg, Germany
17 Association of Physiotherapists in Parkinson’s Disease Europe, Esch-sur-Alzette, Luxembourg
18 Private practice, Ettelbruck, Luxembourg
19 Private practice, Luxembourg, Luxembourg

## COURAGE-PD consortium

Sulev KOKS 1, George D. MELLICK 2, Walter PIRKER 3, Alexander ZIMPRICH 4, Anthony E LANG 5, Ekaterina ROGAEVA 6, Pille TABA 7, Alexis BRICE 8, Marie-Christine CHARTIER-HARLIN 9, Jean-Christophe CORVOL 8, Cloé DOMENIGHETTI 10, Alexis ELBAZ 10, Suzanne LESAGE 8, Eugenie MUTEZ 9, Pierre-Emmanuel SUGIER 10, Ashwin ASHOK KUMAR SREELATHA 11, Sandeep GROVER 11, Kathrin BROCKMANN 12, Angela B. DEUTSCHLANDER 13, Thomas GASSER 12, Jens KRUGER 14, Peter LICHTNER 15, Milena RADIVOJKOV-BLAGOJEVIC 15, Claudia SCHULTE 12, Manu SHARMA 11, Efthimos DARDIOTIS 16, Georges M. HADJIGEORGIOU 17, Athina Maria SIMITSI 18, Leonidas STEFANIS 19, Grazia ANNESI 20, Laura BRIGHINA 21, Carlo FERRARESE 21, Simona PETRUCCI 22, Gianni PEZZOLI 23, Andrea QUATTRONE 24, Letizia STRANIERO 25, Monica GAGLIARDI 26, Enza Maria VALENTE 27, Anna ZECCHINELLI 23, Nobotaka HATTORI 28, Akiyoshi, NAKAYAMA 29, Hirotaka MATSUO 29, Kenya NISHIOKA 28, Dheeraj BOBBILI 30, Rejko KRUGER 31, Zied LANDOULSI 30, Patrick MAY 30, Lukas PAVELKA 31, Bastiaan R. BLOEM 32, Bart P.C. VAN DE WARRENBURG 32, Jan AASLY 33, Lasse PIHLSTROM 34, Mathias TOFT 34, Joaquim J. FERREIRA 35, Leonor CORREIA GUEDES 36, Soraya BARDIEN 37, Jonathan CARR 38, Sun Ju CHUNG 39, Yun Joong KIM 40, Monica DIEZ-FAIREN 41, Mario EZQUERRA 42, Pau PASTOR 41, Eduardo TOLOSA 43, Andrea C. BELIN 44, Nancy L. PEDERSEN 45, Andreas PUSCHMANN 46, Caroline RAN 44, Emil Y. RODSTROM 46, Karin WIRDEFELDT 47, Carl E. CLARKE 48, Karen E. MORRISON 49, Manuela TAN 50, Connor EDSALL 51, Matt J. FARRER 52, Dimitri KRAINC 53, Andrew B. SINGLETON 54, Lena F. BURBULLA 55, Dena G. HERNANDEZ 56.

1 Centre for Molecular Medicine and Innovative Therapeutics, Murdoch University, Murdoch, Australia, Perron Institute for Neurological and Translational Science, Nedlands, Western Australia, Australia.
2 Griffith Institute for Drug Discovery, Griffith University, Don Young Road, Nathan, Queensland, Australia.
3 Department of Neurology, Wilhelminenspital, Austria.
4 Department of Neurology, Medical University of Vienna, Austria.
5 Edmond J. Safra Program in Parkinson’s Disease, Morton and Gloria Shulman Movement Disorders Clinic, Toronto Western Hospital, UHN, Toronto, Ontario, Canada; Division of Neurology, University of Toronto, Toronto, Ontario, Canada; Krembil Brain Institute, Toronto, Ontario, Canada.
6 Tanz Centre for Research in Neurodegenerative Diseases, University of Toronto, Toronto, Ontario, Canada.
7 Department of Neurology and Neurosurgery, University of Tartu, Estonia; Neurology Clinic, Tartu University Hospital, Tartu, Estonia
8 Sorbonne Université, Institut du Cerveau – Paris Brain Institute – ICM, INSERM, CNRS, Assistance Publique Hôpitaux de Paris, Department of Neurology, Paris, France.
9 Univ. Lille, Inserm, CHU Lille, UMR-S 1172 – JPArc – Centre de Recherche Lille Neurosciences & Cognition, F-59000 Lille, France.
10 Université Paris-Saclay, UVSQ, Univ. Paris-Sud, Inserm, Team “Exposome, heredity, cancer and health”, CESP, 94807, Villejuif, France.
11 Centre for Genetic Epidemiology, Institute for Clinical Epidemiology and Applied Biometry, University of Tubingen, Germany.
12 Department for Neurodegenerative Diseases, Hertie Institute for Clinical Brain Research, University of Tubingen, Germany German Center for Neurodegenerative Diseases (DZNE), Tubingen, Germany.
13 Department of Neurology, Ludwig Maximilians University of Munich, Germany; Department of Neurology, Max Planck Institute of Psychiatry, Munich, Germany; Department of Neurology and Department of Clinical Genomics, Mayo Clinic Florida, Jacksonville, FL, USA.
14 Group of Applied Bioinformatics, University of Tubingen, Germany.
15 Institute of Human Genetics, Helmholtz Zentrum München, Neuherberg, Germany.
16 Laboratory of Neurogenetics, Department of Neurology, Faculty of Medicine, University of Thessaly, Larissa, Greece.
17 Department of Neurology, Medical School, University of Cyprus, Nicosia, Cyprus; Department of Neurology, Laboratory of Neurogenetics, University of Thessaly, University Hospital of Larissa, Larissa, Greece.
18 1^st^ Department of Neurology, Eginition Hospital, Medical School, National and Kapodistrian University of Athens, Athens, Greece.
19 Center of Clinical Research, Experimental Surgery and Translational Research, Biomedical Research Foundation of the Academy of Athens, Athens, Greece; 1st Department of Neurology, Eginition Hospital, Medical School, National and Kapodistrian University of Athens, Athens, Greece.
20 Institute for Biomedical Research and Innovation, National Research Council, Cosenza, Italy.
21 Department of Neurology, San Gerardo Hospital, Monza, Italy; Department of Medicine and Surgery and Milan Center for Neuroscience, University of Milano Bicocca, Milano, Italy. 22 Department of Clinical and Molecular Medicine, University of Rome, Italy; UOC Medical Genetics and Advanced Cell Diagnostics, S. Andrea University Hospital, Rome, Italy.
23 Parkinson Institute, Azienda Socio Sanitaria Territoriale (ASST) Gaetano Pini/CTO, Milano, Italy.
24 Institute of Neurology, Magna Graecia University, Catanzaro, Italy.
25 Department of Biomedical Sciences, Humanitas University, Milan, Italy.
26 Institute of Molecular Bioimaging and Physiology National Research Council, Catanzaro, Italy.
27 Department of Molecular Medicine, University of Pavia, Italy; Istituto di Ricovero e Cura a Carattere Scientifico (IRCCS) Mondino Foundation, Pavia, Italy.
28 Department of Neurology, Juntendo University School of Medicine, Bunkyo-ku, Tokyo 113-8421, Japan.
29 Department of Integrative Physiology and Bio-Nano Medicine, National Defense Medical College, Saitama 359-8513, Japan.
30 Bioinformatics Core, Luxembourg Centre for Systems Biomedicine (LCSB), University of Luxembourg, Esch-Belval, Luxembourg.
31 Translational Neuroscience, Luxembourg Centre for Systems Biomedicine (LCSB), University of Luxembourg, Esch-Belval, Luxembourg; Parkinson’s Research Clinic, Centre Hospitalier de Luxembourg, Luxembourg; Transversal Translational Medicine, Luxembourg Institute of Health (LIH), Strassen, Luxembourg.
32 Radboud University Medical Centre, Donders Institute for Brain, Cognition and Behaviour, Department of Neurology, Nijmegen, The Netherlands.
33 Department of Neurology, St Olav’s Hospital and Norwegian University of Science and Technology, Trondheim, Norway.
34 Department of Neurology, Oslo University Hospital, Oslo, Norway.
35 Instituto de Medicina Molecular João Lobo Antunes, Faculdade de Medicina, Universidade de Lisboa, Lisbon, Portugal; Laboratory of Clinical Pharmacology and Therapeutics, Faculdade de Medicina, Universidade de Lisboa, Lisbon, Portugal.
36 Instituto de Medicina Molecular João Lobo Antunes, Faculdade de Medicina, Universidade de Lisboa, Lisbon, Portugal. Campus Neurológico Sénior, Torres Vedras, Portugal; Department of Neurosciences and Mental Health, Neurology, Hospital de Santa Maria, CHULN, Lisbon, Portugal.
37 Division of Molecular Biology and Human Genetics, Department of Biomedical Sciences, Faculty of Medicine and Health Sciences, Stellenbosch University, South Africa; South African Medical Research Council/ Stellenbosch University Genomics of Brain Disorders Research Unit, Cape Town, South Africa.
38 Division of Neurology, Department of Medicine, Faculty of Medicine and Health Sciences, Stellenbosch University, South Africa; South African Medical Research Council/ Stellenbosch University Genomics of Brain Disorders Research Unit, Cape Town, South Africa.
39 Department of Neurology, Asan Medical Center, University of Ulsan College of Medicine, Seoul, South Korea.
40 Department of Neurology, Yongin Severance Hospital, Yonsei University College of Medicine, Seoul, South Korea.
41 Fundació per la Recerca Biomèdica i Social Mútua Terrassa, Terrassa, Barcelona, Spain; Movement Disorders Unit, Department of Neurology, Hospital Universitari Mutua de Terrassa, Terrassa, Barcelona, Spain.
42 Lab of Parkinson Disease and Other Neurodegenerative Movement Disorders, Institut d’Investigacions Biomèdiques August Pi i Sunyer (IDIBAPS), Institut de Neurociències, Universitat de Barcelona, ES-08036 Barcelona, Catalonia, Spain.
43 Parkinson’s disease & Movement Disorders Unit, Neurology Service, Hospital Clínic de Barcelona, Institut d’Investigacions Biomèdiques August Pi i Sunyer (IDIBAPS), University of Barcelona, Barcelona, Spain Centro de Investigación Biomédica en Red sobre Enfermedades Neurodegenerativas (CIBERNED: CB06/05/0018-ISCIII) Barcelona, Spain.
44 Department of Neuroscience, Karolinska Institutet, Stockholm, Sweden.
45 Department of Medical Epidemiology and Biostatistics, Karolinska Institutet, Stockholm, Sweden.
46 Lund University, Skåne University Hospital, Department of Clinical Sciences Lund, Neurology, Getingevägen 4, 221 85, Lund, Sweden.
47 Department of Medical Epidemiology and Biostatistics, Karolinska Institutet, Stockholm, Sweden; Department of Clinical Neuroscience, Karolinska Institutet, Stockholm, Sweden.
48 University of Birmingham and Sandwell and West Birmingham Hospitals NHS Trust, United Kingdom.
49 Faculty of Medicine, Health and Life Sciences, Queens University, Belfast, United Kingdom.
50 Department of Clinical and Movement Neurosciences, UCL Queen Square Institute of Neurology, University College London, London, UK.
51 Molecular Genetics Section, Laboratory of Neurogenetics, NIA, NIH, Bethesda, MD 20892, USA.
52 Department of Neurology, McKnight Brain Institute, University of Florida, Gainesville, FL, USA.
53 Department of Neurology, Northwestern University Feinberg School of Medicine, Chicago, Illinois 60611, United States.
54 Molecular Genetics Section, Laboratory of Neurogenetics, NIA, NIH, Bethesda, MD 20892, USA; Center For Alzheimer’s and Related Dementias, NIA, NIH, Bethesda, MD 20892, USA.
55 Department of Neurology, Northwestern University Feinberg School of Medicine, Chicago, IL 60611, USA; German Center for Neurodegenerative Diseases (DZNE), Munich, Germany; Metabolic Biochemistry, Biomedical Center 13 (BMC), Faculty of Medicine, Ludwig-Maximilians University, Munich, Germany; Munich Cluster for Systems Neurology (SyNergy), Munich, Germany.
56 Molecular Genetics Section, Laboratory of Neurogenetics, NIA, NIH, Bethesda, MD 20892, USA.

## References

1. Ascherio A, Schwarzschild MA. The epidemiology of Parkinson’s disease: risk factors and prevention. The Lancet Neurology. 2016;15:1257–72.

2. Jankovic J, Tan EK. Parkinson’s disease: etiopathogenesis and treatment. J Neurol Neurosurg Psychiatry. 2020;91:795–808.

3. Lesage S, Brice A. Parkinson’s disease: from monogenic forms to genetic susceptibility factors. Human Molecular Genetics. 2009;18:R48–59.

4. Nalls MA, Blauwendraat C, Vallerga CL, Heilbron K, Bandres-Ciga S, Chang D, et al. Identification of novel risk loci, causal insights, and heritable risk for Parkinson’s disease: a meta-analysis of genome-wide association studies. Lancet Neurol. 2019;18:1091–102.

5. Dehestani M, Liu H, Gasser T. Polygenic Risk Scores Contribute to Personalized Medicine of Parkinson’s Disease. JPM. 2021;11:1030.

6. Choi SW, Mak TS-H, O’Reilly PF. Tutorial: a guide to performing polygenic risk score analyses. Nat Protoc. 2020;15:2759–72.

7. Sherer TB, Richardson JR, Testa CM, Seo BB, Panov AV, Yagi T, et al. Mechanism of toxicity of pesticides acting at complex I: relevance to environmental etiologies of Parkinson’s disease. J Neurochem. 2007;0:070214184024016-???

8. González-Rodríguez P, Zampese E, Stout KA, Guzman JN, Ilijic E, Yang B, et al. Disruption of mitochondrial complex I induces progressive parkinsonism. Nature. 2021;599:650–6.

9. Malpartida AB, Williamson M, Narendra DP, Wade-Martins R, Ryan BJ. Mitochondrial Dysfunction and Mitophagy in Parkinson’s Disease: From Mechanism to Therapy. Trends in Biochemical Sciences. 2021;46:329–43.

10. Schapira AHV, Cooper JM, Dexter D, Jenner P, Clark JB, Marsden CD. MITOCHONDRIAL COMPLEX I DEFICIENCY IN PARKINSON’S DISEASE. The Lancet. 1989;333:1269.

11. Angelova PR, Abramov AY. Role of mitochondrial ROS in the brain: from physiology to neurodegeneration. FEBS Lett. 2018;592:692–702.

12. Bender A, Krishnan KJ, Morris CM, Taylor GA, Reeve AK, Perry RH, et al. High levels of mitochondrial DNA deletions in substantia nigra neurons in aging and Parkinson disease. Nat Genet. 2006;38:515–7.

13. Burbulla LF, Krüger R. Converging environmental and genetic pathways in the pathogenesis of Parkinson’s disease. Journal of the Neurological Sciences. 2011;306:1–8.

14. Billingsley KJ, Barbosa IA, Bandrés-Ciga S, Quinn JP, Bubb VJ, Deshpande C, et al. Mitochondria function associated genes contribute to Parkinson’s Disease risk and later age at onset. npj Parkinsons Dis. 2019;5:8.

15. Hipp G, Vaillant M, Diederich NJ, Roomp K, Satagopam VP, Banda P, et al. The Luxembourg Parkinson’s Study: A Comprehensive Approach for Stratification and Early Diagnosis. Front Aging Neurosci. 2018;10:326.

16. Domenighetti C, Sugier P, Ashok Kumar Sreelatha A, Schulte C, Grover S, Mohamed O, et al. Dairy Intake and Parkinson’s Disease: A Mendelian Randomization Study. Movement Disorders. 2022;37:857–64.

17. Grover S, Kumar□Sreelatha AA, Bobbili DR, May P, Domenighetti C, Sugier P, et al. Replication of a Novel Parkinson’s Locus in a European Ancestry Population. Mov Disord. 2021;36:1689–95.

18. Litvan I, Bhatia KP, Burn DJ, Goetz CG, Lang AE, McKeith I, et al. SIC Task Force appraisal of clinical diagnostic criteria for parkinsonian disorders. Mov Disord. 2003;18:467– 86.

19. Blauwendraat C, Faghri F, Pihlstrom L, Geiger JT, Elbaz A, Lesage S, et al. NeuroChip, an updated version of the NeuroX genotyping platform to rapidly screen for variants associated with neurological diseases. Neurobiology of Aging. 2017;57:247.e9–247.e13.

20. Chang CC, Chow CC, Tellier LC, Vattikuti S, Purcell SM, Lee JJ. Second-generation PLINK: rising to the challenge of larger and richer datasets. GigaSci. 2015;4:7.

21. Das S, Forer L, Schönherr S, Sidore C, Locke AE, Kwong A, et al. Next-generation genotype imputation service and methods. Nat Genet. 2016;48:1284–7.

22. Rath S, Sharma R, Gupta R, Ast T, Chan C, Durham TJ, et al. MitoCarta3.0: an updated mitochondrial proteome now with sub-organelle localization and pathway annotations. Nucleic Acids Research. 2021;49:D1541–7.

23. Choi SW, O’Reilly PF. PRSice-2: Polygenic Risk Score software for biobank-scale data. GigaScience. 2019;8:giz082.

24. Cai L, Wheeler E, Kerrison ND, Luan J, Deloukas P, Franks PW, et al. Genome-wide association analysis of type 2 diabetes in the EPIC-InterAct study. Sci Data. 2020;7:393.

25. Bigdeli TB, Fanous AH, Li Y, Rajeevan N, Sayward F, Genovese G, et al. Genome-Wide Association Studies of Schizophrenia and Bipolar Disorder in a Diverse Cohort of US Veterans. Schizophrenia Bulletin. 2021;47:517–29.

26. Kunkle BW, Grenier-Boley B, Sims R, Bis JC, Damotte V, Naj AC, et al. Genetic meta-analysis of diagnosed Alzheimer’s disease identifies new risk loci and implicates Aβ, tau, immunity and lipid processing. Nat Genet. 2019;51:414–30.

27. Reinhardt P, Glatza M, Hemmer K, Tsytsyura Y, Thiel CS, Höing S, et al. Derivation and Expansion Using Only Small Molecules of Human Neural Progenitors for Neurodegenerative Disease Modeling. Daadi M, editor. PLoS ONE. 2013;8:e59252.

28. Jarazo J, Barmpa K, Modamio J, Saraiva C, Sabaté□Soler S, Rosety I, et al. Parkinson’s Disease Phenotypes in Patient Neuronal Cultures and Brain Organoids Improved by 2□HYDROXYPROPYL□Β□CYCLODEXTRIN Treatment. Movement Disorders. 2022;37:80–94.

29. Gu X, Ma Y, Liu Y, Wan Q. Measurement of mitochondrial respiration in adherent cells by Seahorse XF96 Cell Mito Stress Test. STAR Protocols. 2021;2:100245.

30. Swerdlow RH, E. L, Aires D, Lu J. Glycolysis–respiration relationships in a neuroblastoma cell line. Biochimica et Biophysica Acta (BBA) – General Subjects. 2013;1830:2891–8.

31. Maston GA, Evans SK, Green MR. Transcriptional Regulatory Elements in the Human Genome. Annu Rev Genom Hum Genet. 2006;7:29–59.

32. Bandres-Ciga S, Saez-Atienzar S, Kim JJ, Makarious MB, Faghri F, Diez-Fairen M, et al. Large-scale pathway specific polygenic risk and transcriptomic community network analysis identifies novel functional pathways in Parkinson disease. Acta Neuropathol. 2020;140:341– 58.

33. Jung S-H, Kim H-R, Chun MY, Jang H, Cho M, Kim B, et al. Transferability of Alzheimer Disease Polygenic Risk Score Across Populations and Its Association With Alzheimer Disease-Related Phenotypes. JAMA Netw Open. 2022;5:e2247162.

34. Kachuri L, Graff RE, Smith-Byrne K, Meyers TJ, Rashkin SR, Ziv E, et al. Pan-cancer analysis demonstrates that integrating polygenic risk scores with modifiable risk factors improves risk prediction. Nat Commun. 2020;11:6084.

35. Mitchell P. Coupling of Phosphorylation to Electron and Hydrogen Transfer by a Chemi-Osmotic type of Mechanism. Nature. 1961;191:144–8.

36. Chen H, Chomyn A, Chan DC. Disruption of Fusion Results in Mitochondrial Heterogeneity and Dysfunction. Journal of Biological Chemistry. 2005;280:26185–92.

37. Yao C-H, Wang R, Wang Y, Kung C-P, Weber JD, Patti GJ. Mitochondrial fusion supports increased oxidative phosphorylation during cell proliferation. eLife. 2019;8:e41351.

38. Grünewald A, Rygiel KA, Hepplewhite PD, Morris CM, Picard M, Turnbull DM. Mitochondrial DNA Depletion in Respiratory Chain–Deficient Parkinson Disease Neurons. Ann Neurol. 2016;79:366–78.

39. Tang JX, Thompson K, Taylor RW, Oláhová M. Mitochondrial OXPHOS Biogenesis: Co-Regulation of Protein Synthesis, Import, and Assembly Pathways. IJMS. 2020;21:3820.

40. Borsche M, König IR, Delcambre S, Petrucci S, Balck A, Brüggemann N, et al. Mitochondrial damage-associated inflammation highlights biomarkers in PRKN/PINK1 parkinsonism. Brain. 2020;143:3041–51.

41. Sliter DA, Martinez J, Hao L, Chen X, Sun N, Fischer TD, et al. Parkin and PINK1 mitigate STING-induced inflammation. Nature. 2018;561:258–62.

42. Candelario KM, Shuttleworth CW, Cunningham LA. Neural stem/progenitor cells display a low requirement for oxidative metabolism independent of hypoxia inducible factor-1alpha expression. J Neurochem. 2013;125:420–9.

43. Blauwendraat C, Nalls MA, Singleton AB. The genetic architecture of Parkinson’s disease. The Lancet Neurology. 2020;19:170–8.

44. Ohnmacht J, May P, Sinkkonen L, Krüger R. Missing heritability in Parkinson’s disease: the emerging role of non-coding genetic variation. J Neural Transm. 2020;127:729–48.

45. Dehestani M, Liu H, Sreelatha AAK, Schulte C, Bansal V, Gasser T. Mitochondrial and autophagy-lysosomal pathway polygenic risk scores predict Parkinson’s disease. Molecular and Cellular Neuroscience. 2022;121:103751.

46. Paliwal D, McInerney TW, Pa J, Swerdlow RH, Easteal S, Andrews SJ. Mitochondrial pathway polygenic risk scores are associated with Alzheimer’s Disease. Neurobiology of Aging. 2021;108:213–22.

47. Khera AV, Chaffin M, Aragam KG, Haas ME, Roselli C, Choi SH, et al. Genome-wide polygenic scores for common diseases identify individuals with risk equivalent to monogenic mutations. Nat Genet. 2018;50:1219–24.

48. Landi I, Kaji DA, Cotter L, Van Vleck T, Belbin G, Preuss M, et al. Prognostic value of polygenic risk scores for adults with psychosis. Nat Med. 2021;27:1576–81.

49. Leonenko G, Baker E, Stevenson-Hoare J, Sierksma A, Fiers M, Williams J, et al. Identifying individuals with high risk of Alzheimer’s disease using polygenic risk scores. Nat Commun. 2021;12:4506.

50. Adeyemo A, Balaconis MK, Darnes DR, Fatumo S, Granados Moreno P, Hodonsky CJ, et al. Responsible use of polygenic risk scores in the clinic: potential benefits, risks and gaps. Nat Med. 2021;27:1876–84.

51. Mor DE, Sohrabi S, Kaletsky R, Keyes W, Tartici A, Kalia V, et al. Metformin rescues Parkinson’s disease phenotypes caused by hyperactive mitochondria. Proc Natl Acad Sci USA. 2020;117:26438–47.

52. Mor DE, Murphy CT. Mitochondrial hyperactivity as a potential therapeutic target in Parkinson’s disease. Translational Medicine of Aging. 2020;4:117–20.

53. Keeney PM. Parkinson’s Disease Brain Mitochondrial Complex I Has Oxidatively Damaged Subunits and Is Functionally Impaired and Misassembled. Journal of Neuroscience. 2006;26:5256–64.

54. Dias V, Junn E, Mouradian MM. The Role of Oxidative Stress in Parkinson’s Disease. Journal of Parkinson’s Disease. 2013;3:461–91.

55. Haylett W, Swart C, van der Westhuizen F, van Dyk H, van der Merwe L, van der Merwe C, et al. Altered Mitochondrial Respiration and Other Features of Mitochondrial Function in *Parkin*-Mutant Fibroblasts from Parkinson’s Disease Patients. Parkinson’s Disease. 2016;2016:1–11.

56. Zanellati MC, Monti V, Barzaghi C, Reale C, Nardocci N, Albanese A, et al. Mitochondrial dysfunction in Parkinson disease: evidence in mutant PARK2 fibroblasts. Front Genet [Internet]. 2015 [cited 2022 Jun 29];6. Available from: http://www.frontiersin.org/Genetic_Disorders/10.3389/fgene.2015.00078/abstract

57. Annesley SJ, Lay ST, De Piazza SW, Sanislav O, Hammersley E, Allan CY, et al. Immortalized Parkinson’s Disease lymphocytes have enhanced mitochondrial respiratory activity. Disease Models & Mechanisms. 2016;dmm.025684.

58. Auburger G, Klinkenberg M, Drost J, Marcus K, Morales-Gordo B, Kunz WS, et al. Primary Skin Fibroblasts as a Model of Parkinson’s Disease. Mol Neurobiol. 2012;46:20–7.

59. Kim D-Y, Rhee I, Paik J. Metabolic circuits in neural stem cells. Cell Mol Life Sci. 2014;71:4221–41.

60. Chaturvedi RK, Beal MF. Mitochondrial Approaches for Neuroprotection. Annals of the New York Academy of Sciences. 2008;1147:395–412.

61. The Parkinson Study Group QE3 Investigators, Beal MF, Oakes D, Shoulson I, Henchcliffe C, Galpern WR, et al. A Randomized Clinical Trial of High-Dosage Coenzyme Q10 in Early Parkinson Disease: No Evidence of Benefit. JAMA Neurol. 2014;71:543.

62. Prasuhn J, Kasten M, Vos M, König IR, Schmid SM, Wilms B, et al. The Use of Vitamin K2 in Patients With Parkinson’s Disease and Mitochondrial Dysfunction (PD-K2): A Theranostic Pilot Study in a Placebo-Controlled Parallel Group Design. Front Neurol. 2021;11:592104.

63. Kalyanaraman B. Teaching the basics of repurposing mitochondria-targeted drugs: From Parkinson’s disease to cancer and back to Parkinson’s disease. Redox Biology. 2020;36:101665.

64. Carling PJ, Mortiboys H, Green C, Mihaylov S, Sandor C, Schwartzentruber A, et al. Deep phenotyping of peripheral tissue facilitates mechanistic disease stratification in sporadic Parkinson’s disease. Progress in Neurobiology. 2020;187:101772.

65. Antony PMA, Kondratyeva O, Mommaerts K, Ostaszewski M, Sokolowska K, Baumuratov AS, et al. Fibroblast mitochondria in idiopathic Parkinson’s disease display morphological changes and enhanced resistance to depolarization. Sci Rep. 2020;10:1569.

