## Supplementary informations for "Polygenic risk scores validated in patient-derived cells stratify for mitochondrial subtypes of Parkinson’s disease"

**List of Supplementary Materials**

**Materials and Methods**

- ROS measurements
- Analysis of mitochondrial morphology and membrane potential
- Analysis of electron transport chain subunits expression
- Analysis mtDNA copy number, transcription/replication and deletions
- Quantification of plasma IL-6
- RT-qPCR
- Immunocytochemistry
- Karyotype and cell identity analysis

**Supplementary Figures**

- Supplementary Fig. 1
- Supplementary Fig. 2
- Supplementary Fig. 3
- Supplementary Fig. 4
- Supplementary Fig. 5
- Supplementary Fig. 6
- Supplementary Fig. 7
- Supplementary Fig. 8
- Supplementary Fig. 9
- Supplementary Fig. 10
- Supplementary Fig. 11

**Supplementary Tables**

- Supplementary Table 1
- Supplementary Table 2
- Supplementary Table 3
- Supplementary Table 4

**References**

**Materials and Methods**

**ROS measurements**

Primary skin fibroblasts (10000 cells per well) were seeded into Cell Carrier Ultra 96-well plates (Perkin Elmer, #6055302). The next day, fibroblasts medium was removed and replaced by fresh medium containing either 5µM CellROX^TM^ Deep Red reagent (Thermo Fisher Scientific, #C10422) or vehicle only (DMSO). 1µg/ml Hoechst 33342 (Thermo Fisher Scientific, #H21492) was added to both solutions to stain the nuclei. After 30 minutes incubation at 37°C in the dark, cells were washed twice in PBS before proceeding with confocal microscopy analysis. Z-stack images were acquired at 20x magnification using the CellVoyager CV8000 High-Content Screening System (Yokogawa). An in-house Matlab image processing pipeline was used to quantify the CellROX signal. CellROX-positive structures were segmented by preprocessing of the CellROX channel via a Difference of Gaussian (DoG) filter with side-length 110 pixels and sigmas of 1 and 33 pixels, prior to thresholding (>10). CellROX total raw intensities within this mask where normalized to nuclear area to extract numeric features for each field. Nine fields per well and at least two wells per condition were assessed for each of the three independent biological replicates.

**Analysis of mitochondrial morphology and membrane potential**

Primary skin fibroblasts (10000 cells per well) were seeded into Cell Carrier Ultra 96-well plates (Perkin Elmer, #6055302). The next day, fibroblast medium was removed and replaced by FBS-deprived DMEM containing vehicle (DMSO) or 100nM Tetramethylrhodamine, Ethyl Ester, Perchlorate (TMRE, Thermo Fisher Scientific, #T669), a fluorescent dye that specifically accumulates in active, polarized mitochondria

The mitochondrial OXPHOS uncoupler carbonyl cyanide 3-chlorophenylhydrazone (CCCP, Abcam, #ab141229) was used – at the concentration of 50µM – as a positive control of TMRE staining. Nuclei were stained with 1µg/ml Hoechst 33342 (Thermo Fisher Scientific, #H21492). Cells were incubated in the dark at 37°C for 30 minutes before proceeding with confocal microscopy analysis. Z-stack images were acquired at 20x magnification using the CellVoyager CV8000 High-Content Screening System (Yokogawa). An in-house Matlab image processing pipeline was used to quantify the TMRE signal. The mitochondria channel was sum-projected on the x-y plane and preprocessed using a DoG filter of 33 pixel side-length, and sigmas set to 1 and 1.5 pixels, prior to thresholding (>10). Connected components with less than 6 pixels were removed from mitochondrial analysis. Total mitochondrial TMRE fluorescence intensity per field was normalized to nuclear area. Morphometric analysis of the mitochondrial network was performed as described previously [1]. Nine fields per well and at least two wells per condition were assessed for each of the three independent biological replicates.

**Analysis of electron transport chain subunits expression**

Primary skin fibroblasts were washed twice in Phosphate-Buffered Saline (PBS) and then lysed with 1% sodium dodecyl sulfate (SDS) supplemented with Complete Mini EDTA-free Proteinase Inhibitor Cocktail (Merck, #11836170001). Cell lysates were boiled for 5 minutes at 95°C, sonicated and protein quantification performed using the Pierce^TM^ BCA protein assay (Thermo Fisher Scientific, #23227). 10µg of protein per sample were first separated by SDS-PAGE (NUPAGE^TM^, Thermo Fisher Scientific) and then transferred onto a nitrocellulose membrane. The latter was incubated for 1 hour in 5% (w/v) nonfat-dried milk dissolved in TBS-T buffer (10mM Tris-HCl pH 8.0, 150mM NaCl and 0.05% Tween 20), followed by overnight incubation at 4°C with the Total OXPHOS Human WB Antibody Cocktail (Abcam, ab110411). The membrane was washed in TBS-T for 30 minutes and then incubated for 1 hour with a peroxidase (HRP)-conjugated anti-mouse secondary antibody. After 30 minutes of washing in TBS-T, the membrane was incubated with the ECL^TM^ Prime Western Blotting detection reagent (Merck, #GERPN2232) and chemiluminescent visualization of protein bands was performed on the STELLA Bio-imaging system (Raytest Isotopenmessgeräte GmbH, [www.raytest.com](http://www.raytest.com)). After removal of conjugated antibodies by means of the Restore^TM^ PLUS Western Blot Stripping Buffer (Thermo Fisher Scientific, #46430), the same blotting membrane was incubated with a β-actin primary antibody (Cell signaling, #3700), as a control of protein loading.

**Analysis mtDNA copy number, transcription/replication and deletions**

gDNA was extracted from fibroblasts using the QIAmp DNA mini kit (Qiagen, #51304) following the manufacturer’s instructions. Measurement of mtDNA copy number was performed by targeting the mitochondrial gene *ND1* and the nuclear single-copy gene *B2M* using a digital PCR approach (QuantStudio 3D Digital PCR System, Applied Biosystems) [2]. 7S DNA as well as mtDNA major arc deletions were assessed using previously published methods by multiplex qPCR [3,4].

**Quantification of plasma IL-6**

IL-6 levels were measured in plasma samples from the highest or lowest *OXPHOS*-PRS groups using the IL-6 high-sensitive ELISA kit (Enzo, #ENZ-KIT 178-0001), following the manufacturer’s instructions.

**RT-qPCR**

Total RNA was extracted using the RNeasy plus mini kit (Qiagen, #74136), after homogenization of the cell lysate by means of the QIAshredder spin columns (Qiagen, #79654). 1µg of RNA was retrotranscribed using the High-capacity cDNA Reverse Transcription kit (Thermo Fisher Scientific, #4368814). The resulting cDNAs were used to quantify the pluripotency markers *OCT3/4* and *NANOG* by multiplex qPCR, using the LightCycler® 480 Probes Master kit (Roche, #04707494001) combined with the following Taqman probes: *OCT3/4*-FAM (Thermo Fisher Scientific, Hs00999632_g1), NANOG-FAM (Thermo Fisher Scientific, Hs02387400_g1) and ACTB-VIC (Thermo Fisher Scientific, Hs03023880_g1).

**Immunocytochemistry**

iPSCs were seeded in 24-well plates containing Geltrex-coated glass coverslips. Three days after seeding, iPSC medium was removed, followed by two washes in PBS and fixation in 4% paraformaldehyde (PFA). After 15 minutes of incubation, PFA was removed and the coverslips washed twice in PBS before proceeding with the standard immunofluorescence protocol. Briefly, cells were incubated for 1 hour in permeabilization and blocking solution (i.e., PBS containing 0.4% Triton-X100, 10% normal goat serum and 2% BSA), followed by two washes in PBS. Cells were then incubated overnight at 4°C with a rabbit anti-NANOG primary antibody (Abcam, #ab21624), diluted 1:250 in PBS containing 0.1% Triton-X100, 1% normal goat serum and 0.2% BSA. The following day, cells were washed three times in PBS and then incubated for 2 hours (at room temperature) with a goat anti-Rabbit Alexa Fluor 488-conjugated secondary antibody (Thermo Fisher Scientific, #A11008), diluted 1:1000 in PBS containing 0.1% Triton-X100, 1% normal goat serum and 0.2% BSA. After three washes in PBS, nuclei were stained with 1µg/ml Hoechst 33342 (Thermo Fisher Scientific, #H21492) and glass coverslips finally mounted onto the slide in the presence of 10µl VECTASHIELD Antifade Mounting medium (Vector Laboratories, #H1000). Images were acquired using a Zeiss spinning disk confocal microscope (Carl Zeiss Microimaging GmbH).

**Karyotype and cell identity analysis**

Genomic DNA (gDNA) was purified from frozen iPSCs pellets according to the Genomic DNA Purification kit (Thermo Fisher Scientific, #K0512). 100ng total gDNA was used to prepare the Cytoscan HT-CMA 96 array for KaryoStat+ analysis (Thermo Fisher Scientific), based on copy number variants (CNV) and SNPs optimized for balanced whole-genome coverage. The KaryoStat+ assay enables the detection of chromosomal aberrations, including aneuploidies, submicroscopic aberrations and mosaic events, with a resolution similar to g-banding karyotyping (> 1Mb for chromosomal gains and losses). Using the same array as the KaryoStat assay, the cell identity (Cell ID) assay allows for DNA fingerprint matching between newly-generated iPSC lines and the original fibroblast lines, through correlation analysis of 150k SNP probes across the genome of both cell types. This approach enables to detect unique DNA based signatures of the genetic background of a cell, which can then be used for comparative analysis.

**Supplementary Figures**

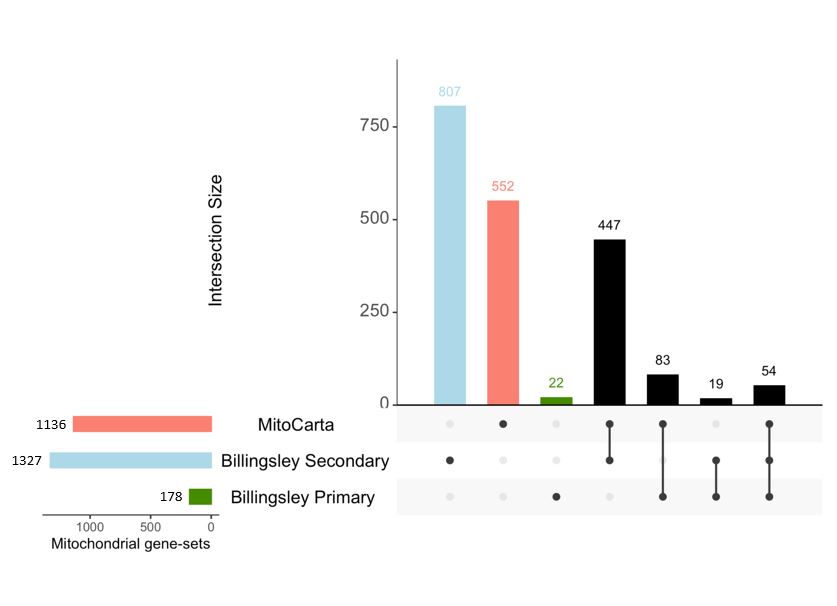

**Supplementary Fig. 1:** **Overlap between mitochondrial genes sets.** Upset plot showing the size (left) and the intersection (right) of three mitochondrial gene sets: Human MitoCarta 3.0, Billingsley Primary and Billingsley Secondary, the latter taken from Billingsley et al [5].

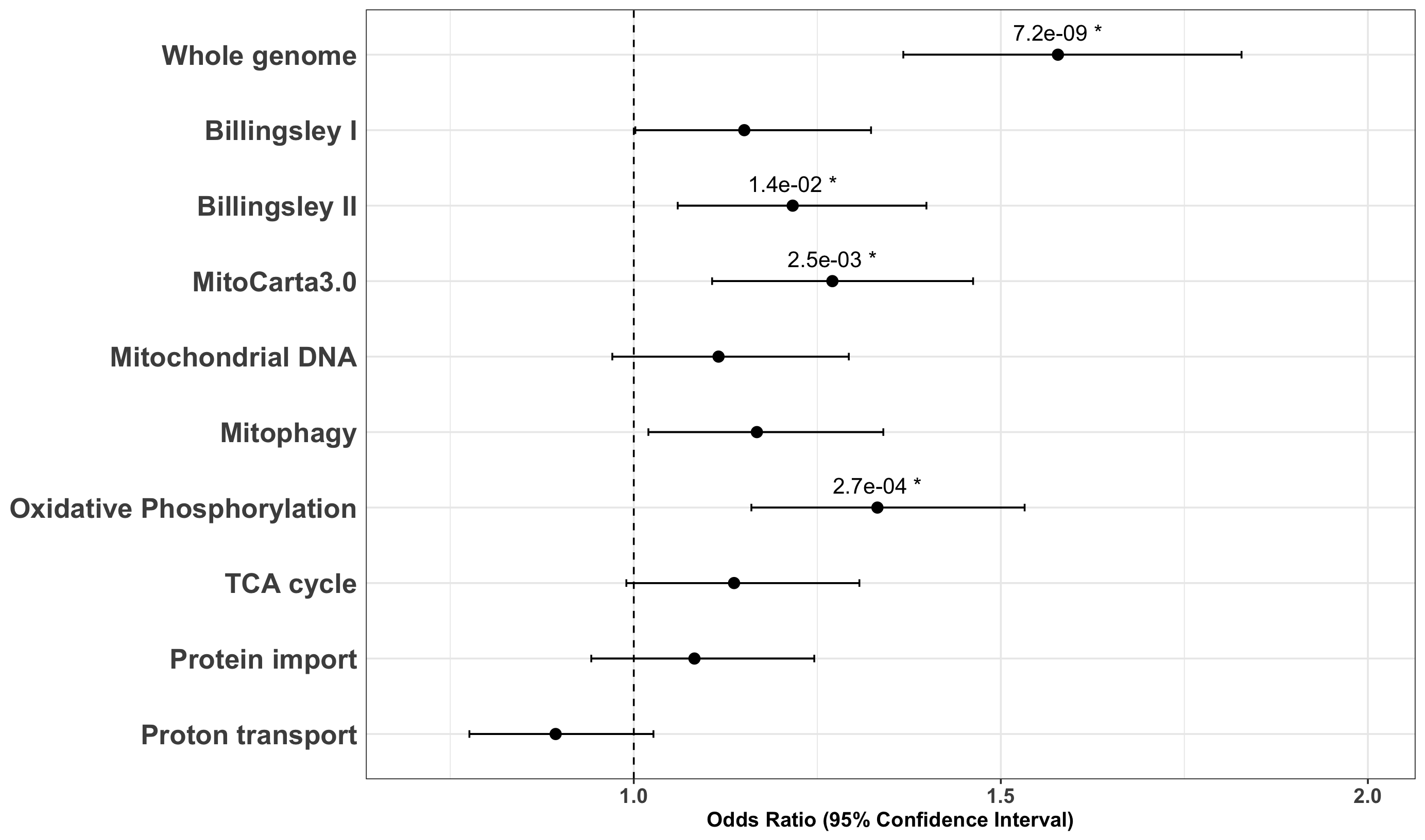

**Supplementary Fig. 2: Association between mitoPRS and PD risk including SNPs in gene regulatory elements.** Forest plots of the odds ratio and 95% confidence interval of polygenic risk scores (PRS) regressed with PD diagnosis in the Luxembourg Parkinson’s study for the whole genome and nine mitochondrial gene sets. Gene boundaries were extended to potential regulatory elements by adding a window of 35kb upstream and 10kb downstream of each gene. * *p* < 0.05.

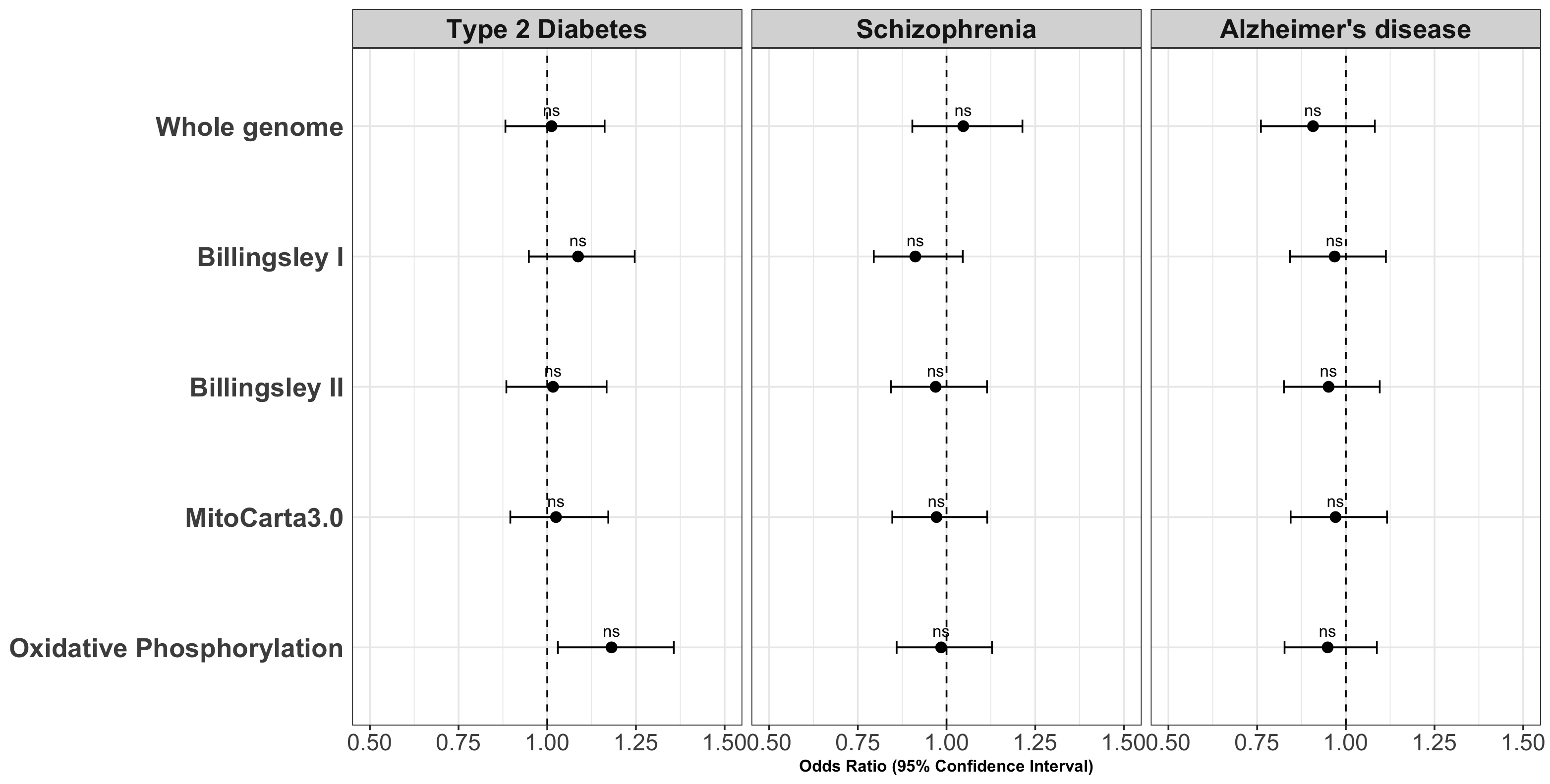

**Supplementary Fig. 3:** **Common risk alleles for other diseases are not associated with higher PD risk in the Luxembourg Parkinson’s Study.** Forest plots of the odds ratio and 95% confidence interval of polygenic risk scores (PRS) regressed with PD diagnosis for Luxembourg Parkinson’s Study, calculated based on Type 2 Diabetes, Schizophrenia and Alzheimer’s disease genome-wide association studies. ns = non-significant *p* value.

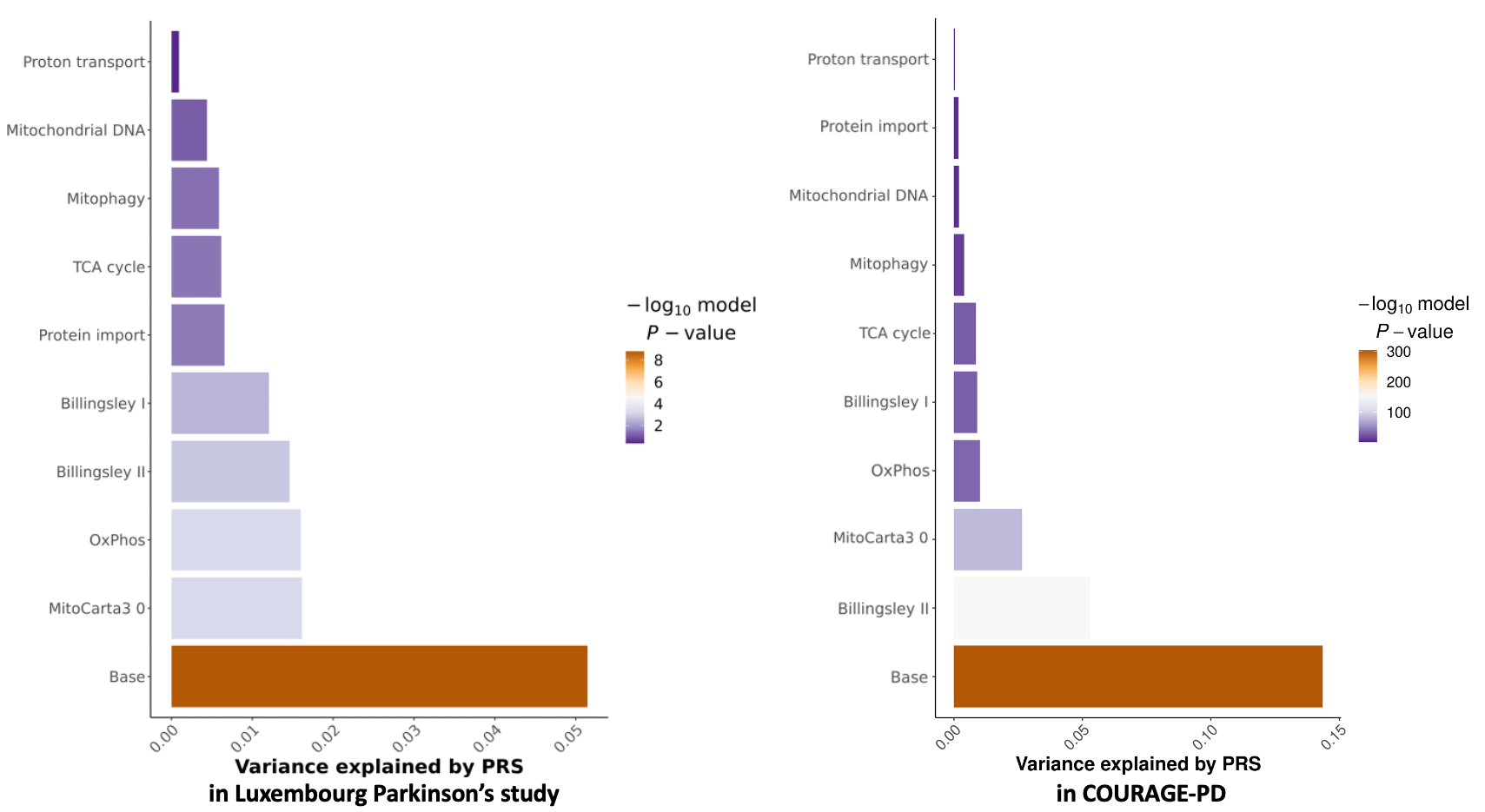

**Supplementary Fig. 4: Phenotypic variance of the mitoPRS models.** Multi-Set plots generated by PRSice2 illustrate the phenotypic variance explained by the PD polygenic risk scores for mitochondrial gene sets and pathways, both for the Luxembourg Parkinson’s study (left panel) and for the COURAGE-PD (right panel) cohorts.

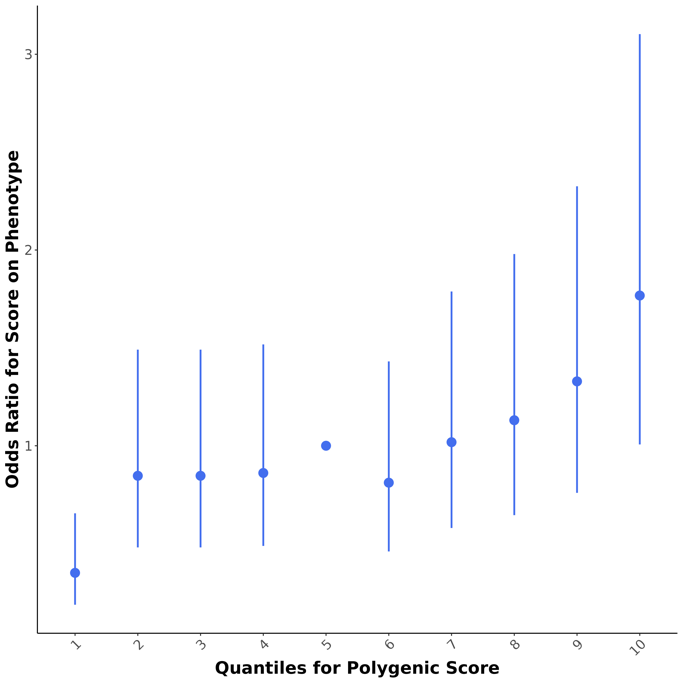

**Supplementary Fig. 5: Quantile plots.** Quantile plots generated by PRSice2 illustrate the fold change in genetic risk across the Luxembourg Parkinson’s study cohort by plotting the odds ratio of PD risk and the polygenic score deciles.

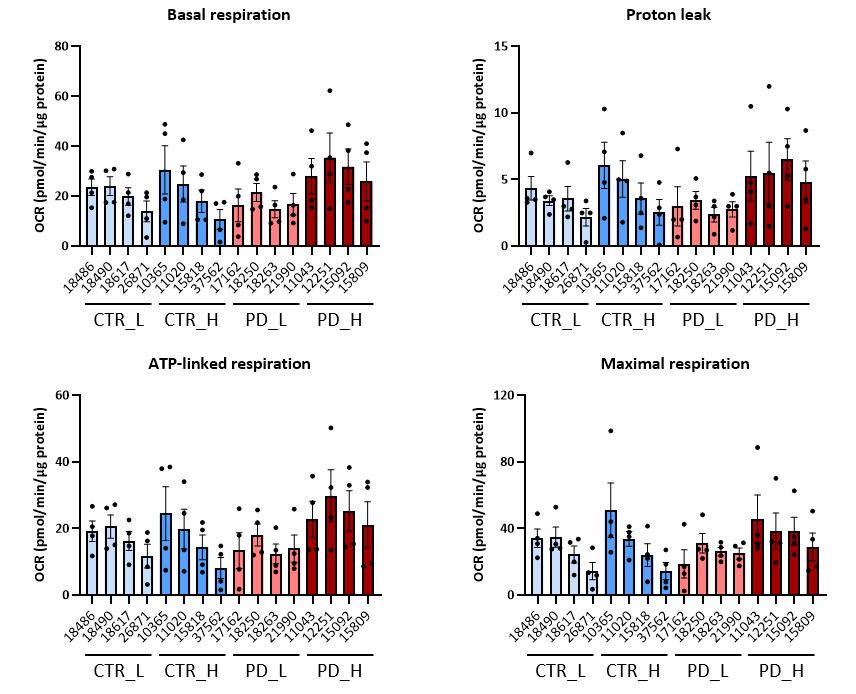

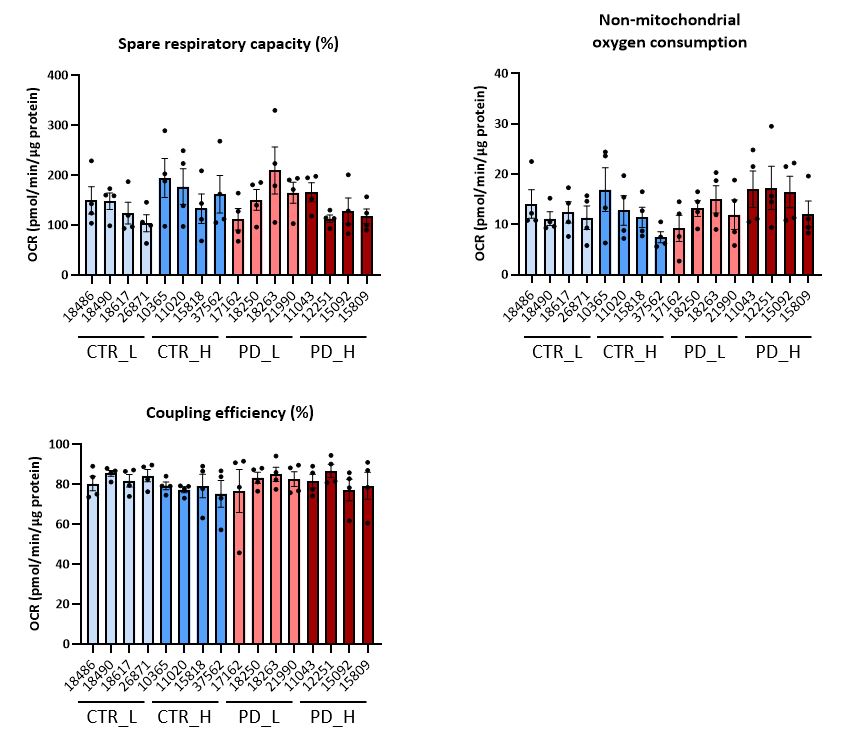

**Supplementary Fig. 6: Analysis of mitochondrial respiration in primary skin fibroblasts from iPD patients and HC stratified based on *OXPHOS*-PRS.** Oxygen consumption rates (OCRs) were measured under basal conditions and after targeted inhibition of specific respiratory chain complexes by using a standard Seahorse Mito Stress test. Histobars represent the means of four independent experiments performed in four distinct fibroblast lines established from Luxembourg Parkinson’s Study participants (healthy controls vs iPD patients) with high (CTR_H ; PD_H) or low (CTR_L ; PD_L) *OXPHOS*-PRS.

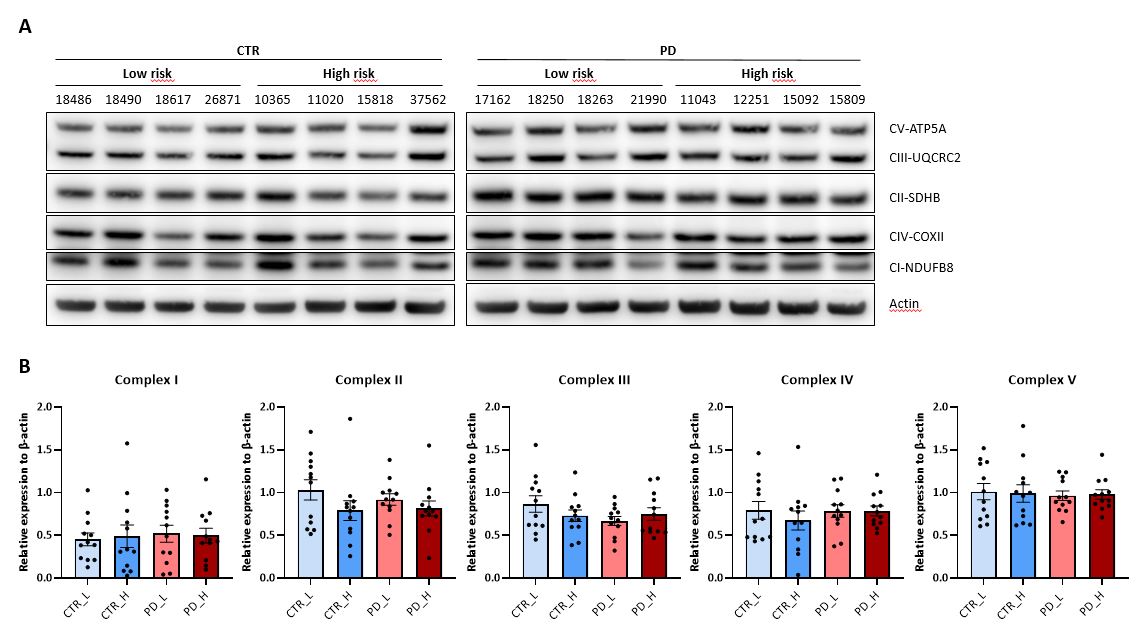

**Supplementary Fig. 7: Analysis of mitochondrial electron transport chain subunits expression in primary skin fibroblasts from iPD patients and HC stratified based on *OXPHOS*-PRS.** (**A**) Representative immunoblots of mitochondrial electron transport chain (ETC) subunits protein expression. β-actin was used as loading control. (**B**) Quantitative densitometric analysis of immunoblotting experiments as in (A). Histobars represent the pooled means of three independent experiments performed in four distinct fibroblast lines established from Luxembourg Parkinson’s Study participants (healthy controls vs iPD patients) with high (CTR_H ; PD_H) or low (CTR_L ; PD_L) *OXPHOS*-PRS. Expression of ETC subunits was normalized against β-actin protein levels.

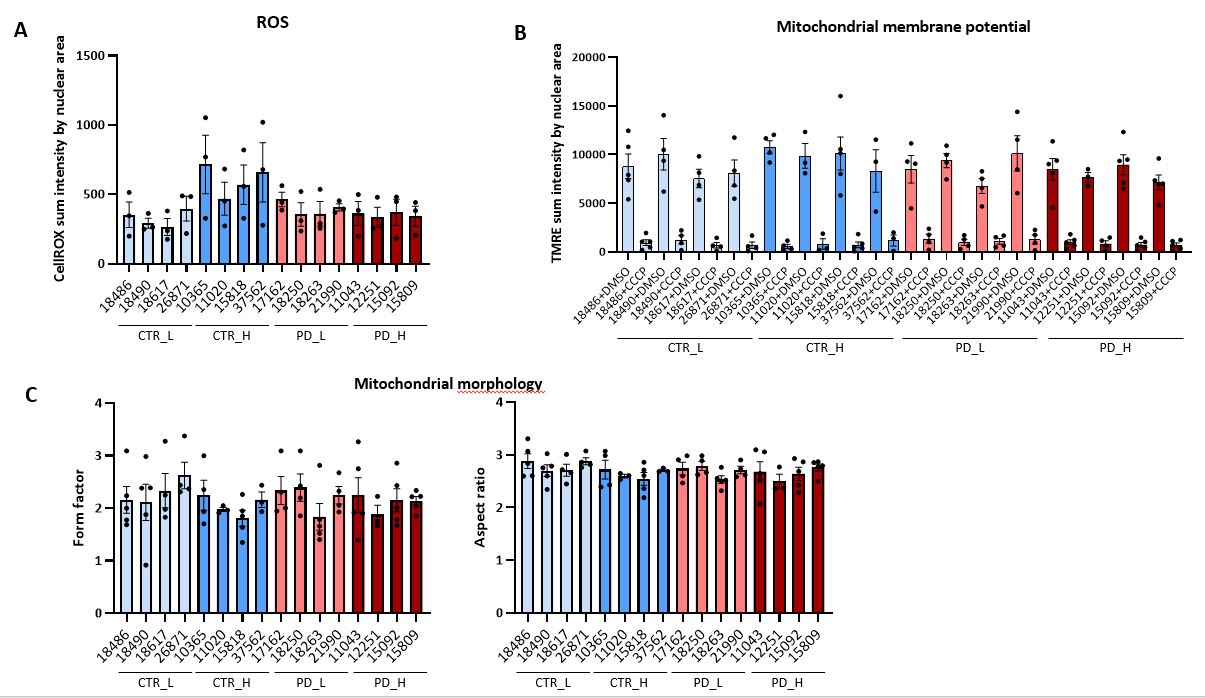

**Supplementary Fig. 8: Confocal microscopy analyses in primary skin fibroblasts from iPD patients and HC stratified based on *OXPHOS*-PRS.** (**A-C**) High-throughput confocal microscopy analyses in primary skin fibroblasts derived from Luxembourg Parkinson’s Study participants (healthy controls vs iPD patients) with high (CTR_H ; PD_H) or low (CTR_L ; PD_L) *OXPHOS*-PRS. (**A**) ROS levels were quantified by normalizing the CellRox mean fluorescence intensity against the nuclear area, as defined by the Hoechst staining. Histobars represent the means of three independent experiments performed in four distinct fibroblast lines for each group. (**B**) Mitochondrial membrane potential (∆Ψ_m_) was measured after normalization of TMRE mean fluorescence intensity by the nuclear area. Treatment with the OXPHOS uncoupler carbonyl cyanide 3-chlorophenylhydrazone (CCCP), known to induce mitochondrial depolarization, was used as positive control for decreased ∆Ψ_m_. Dimethyl sulfoxide (DMSO) was used as a vehicle. Histobars represent the means of at least three independent experiments performed in four distinct fibroblast lines for each group. (**C**) Morphometric analysis of the mitochondrial network. *Form factor* and *aspect ratio* were quantified as described previously *(1)*. Histobars represent the means of at least three independent experiments performed in four distinct fibroblast lines for each group.

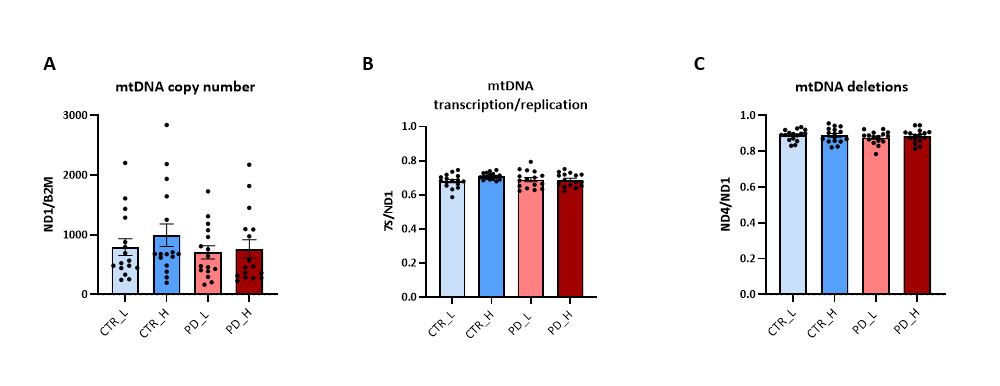

**Supplementary Fig. 9: Assessment of mtDNA integrity in primary skin fibroblasts from iPD patients and HC stratified based on *OXPHOS*-PRS.** (**A-C**) Assessment of mtDNA integrity by triplex qPCR assay. (**A**) mtDNA copy number was assessed by calculating the ratio between *Mitochondrially Encoded NADH:Ubiquinone Oxidoreductase Core Subunit 1 (ND1)* and *β2-microglobulin (B2M)*. (**B**) mtDNA transcription/replication was assessed by measuring the ratio between *7S* DNA and *ND1*. (**C**) Deletions in the major arc of the mitochondrial genome were assessed by calculating the ratio between *Mitochondrially Encoded NADH-dehydrogenase 4 (ND4)* and *ND1*. Histobars represent the pooled means of four independent experiments performed in four distinct fibroblast lines derived from Luxembourg Parkinson’s Study participants (healthy controls vs iPD patients) with high (CTR_H ; PD_H) or low (CTR_L ; PD_L) *OXPHOS*-PRS.

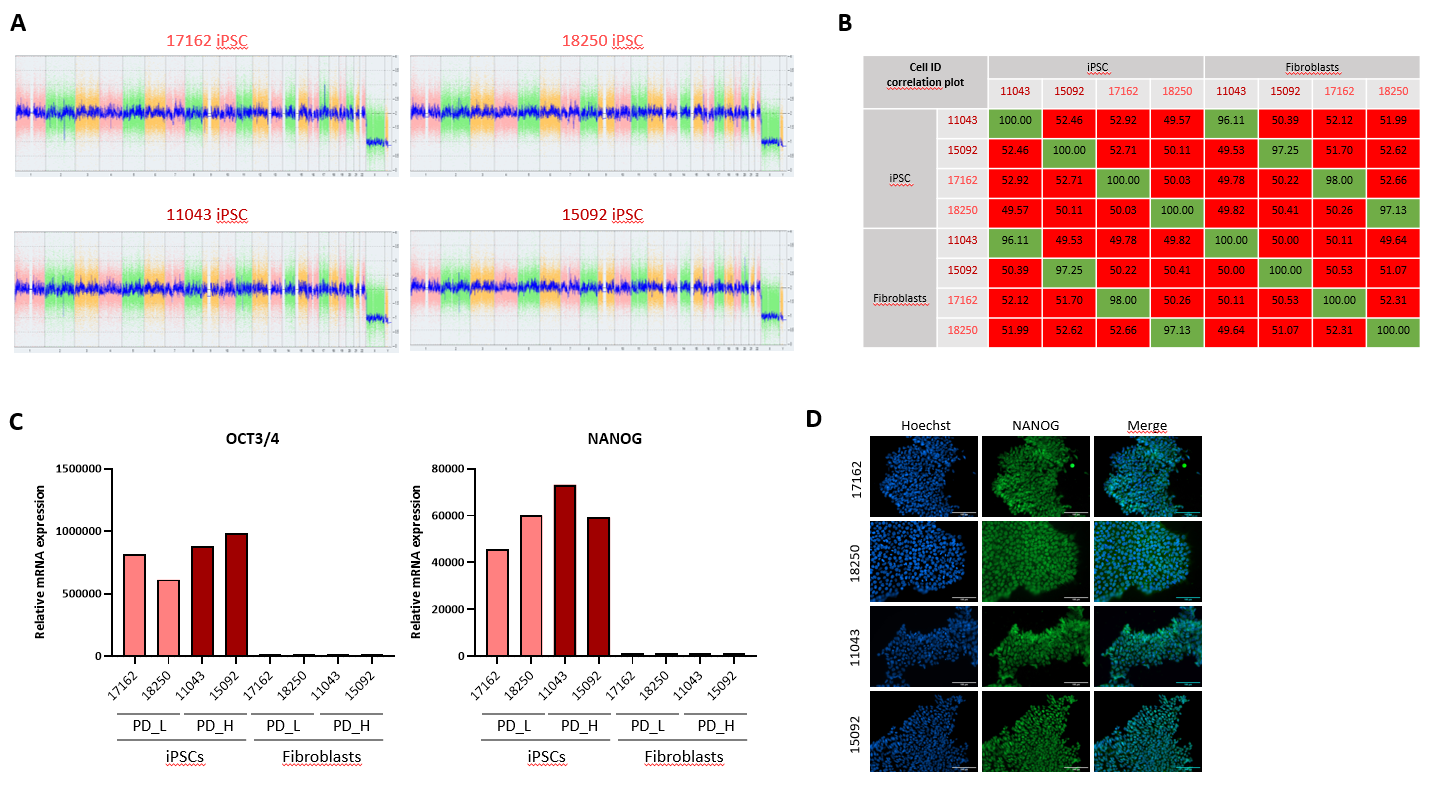

**Supplementary Fig. 10: Characterization of iPSC lines established from iPD patients’ fibroblasts with high or low *OXPHOS*-PRS.** (**A**) Array-based karyotyping of iPSC lines. Somatic and sex chromosomes are represented in the whole genome view. The smooth signal plot (right y-axis) is the smoothing of the log2 ratios which depict the signal intensities of probes on the microarray. A value of 2 represents a normal copy number state (CN = 2). A value of 3 represents chromosomal gain (CN = 3). A value of 1 represents a chromosomal loss (CN = 1). The pink, green and yellow colors indicate the raw signal for each individual chromosome probe, while the blue signal represents the normalized probe signal which is used to identify copy number and aberrations. (**B**) Correlation plot showing DNA fingerprint-based matching of iPSC and fibroblast lines. 150k single nucleotide polymorphisms (SNPs) spread across the genome of both cell types were examined, followed by a correlation analysis of all SNP calls. The correlation between a sample and itself has a value of 100, correlations >95% between samples are considered having an identical genetic background (green boxes), correlations <95% indicate different genetic backgrounds (red boxes). (**C**) RT-qPCR analysis of the stemness markers *OCT3/4* and *NANOG* in iPSCs and primary skin fibroblasts. *OCT3/4* and *NANOG* mRNA expression levels were normalized against *ACTB*. (**D**) Immunofluorescence analysis of the stem cell-specific transcription factor NANOG in the newly generated iPSC lines. Cells expressing NANOG were stained in green. Nuclei were stained with Hoechst. Scale bar: 100µm.

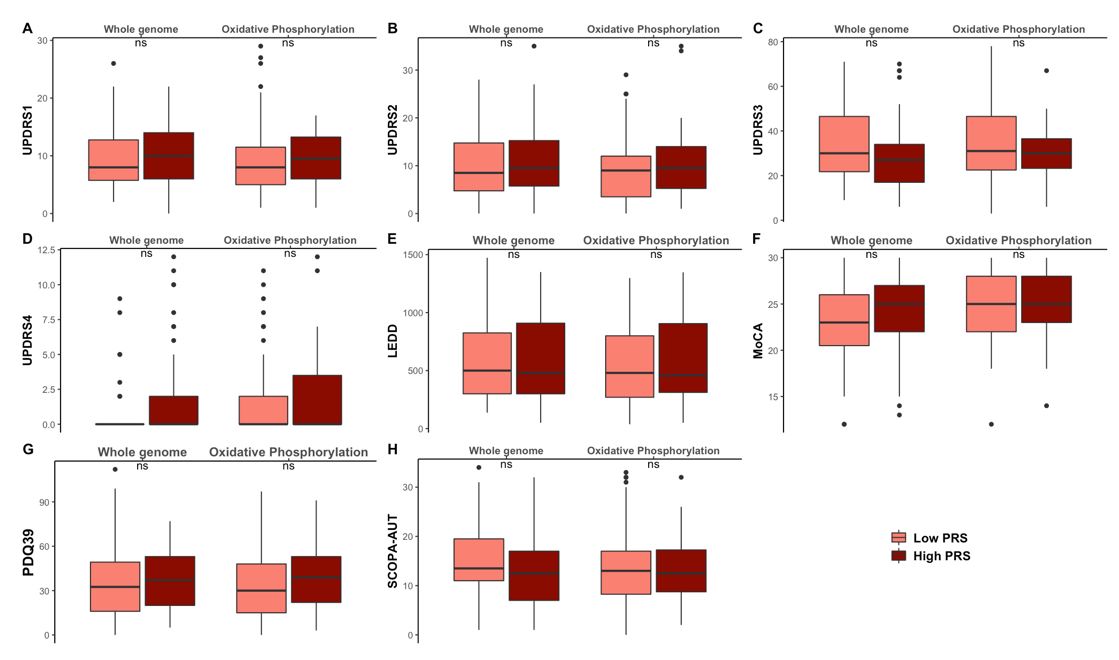

**Supplementary Fig. 11:** **Clinical scores in iPD patients from the Luxembourg Parkinson’s Study with high or low *OXPHOS*-PRS.** Score distributions of MDS-UPDRS-I, II, III, IV, LEDD, MoCA, PDQ39, and SCOPA-AUT were plotted for iPD patients with high (90%-100%) and low (0-10%) *OXPHOS*-PRS. Whole genome PRS was used as a reference. ns = non-significant *p* value.

**Supplementary Tables**

**Supplementary Table 1: List of genes used for the calculation of pathway-specific mitoPRSs.**

| **Gene-sets** | **Pathways ID** | **Genes** |
| --- | --- | --- |
| **Mitophagy**  **(n = 45)** | **GO:0000423** Mitophagy  **GO:1901524** Regulation of mitophagy  **R-HSA-5205647** Reactome mitophagy | *AMBRA1, ATG13, ATG14, BECN1, CDC37, CLEC16A, HDAC6, HTRA2, MFN2, OPTN, PINK1, PRKN, RNF41, SMURF1, SQSTM1, TIGAR, TOMM7, TSC2, USP30, VPS13C, VPS13D, ATG12, ATG5, CSNK2A1, CSNK2A2, CSNK2B, FUNDC1, MAP1LC3A, MAP1LC3B, MFN1, RPS27A, MTERF3, PGAM5, SRC, TOMM20, TOMM22, TOMM40, TOMM5, TOMM6, TOMM70, UBA52, UBB, UBC, ULK1, VDAC1* |
| **TCA cycle**  **(n = 177)** | **GO:0006099** Tricarboxylic acid cycle  **GO:0030062** Mitochondrial tricarboxylic acid cycle enzyme complex  **GO:0045239** Tricarboxylic acid cycle enzyme complex  **REACT_111083** Reactome tca cycle and respiratory electron transport  **R-HSA-1428517** Reactome the citric acid tca cycle and respiratory electron transport | *ACO1, ACO2, CS, DHTKD1, DLAT, DLD, DLST, FAHD1, FH, IDH1, IDH2, IDH3A, IDH3B, IDH3G, IREB2, MDH1, MDH1B, MDH2, ME2, ME3, NNT, OGDH, OGDHL, PDHA1, PDHA2, PDHB, SDHA, SDHAF2, SDHB, SDHC, SDHD, SUCLA2, SUCLG1, SUCLG2, BCKDHA, BCKDHB, BCKDK, DBT, MRPS36, KAT2A, ADHFE1, ATP5F1A, ATP5F1B, ATP5F1C, ATP5F1D, ATP5F1E, ATP5MC1, ATP5ME, ATP5MF, ATP5MG, ATP5PB, ATP5PD, ATP5PF, ATP5PO, BSG, COX4I1, COX5A, COX5B, COX6A1, COX6B1, COX6C, COX7A2L, COX7B, COX7C, COX8A, CYC1, CYCS, D2HGDH, ETFA, ETFB, ETFDH, L2HGDH, LDHA, LDHB, NDUFA1, NDUFA10, NDUFA11, NDUFA12, NDUFA13, NDUFA2, NDUFA3, NDUFA4, NDUFA5, NDUFA6, NDUFA7, NDUFA8, NDUFA9, NDUFAB1, NDUFB1, NDUFB10, NDUFB2, NDUFB3, NDUFB4, NDUFB5, NDUFB6, NDUFB7, NDUFB8, NDUFB9, NDUFC1, NDUFC2, NDUFS1, NDUFS2, NDUFS3, NDUFS4, NDUFS5, NDUFS6, NDUFS7, NDUFS8, NDUFV1, NDUFV2, NDUFV3, PDHX, PDK1, PDK2, PDK3, PDK4, PDP1, PDP2, PDPR, SLC16A1, SLC16A3, SLC16A8, UCP1, UCP2, UCP3, UQCR11, UQCRB, UQCRBP1, UQCRC1, UQCRC2, UQCRFS1, UQCRH, UQCRHL, UQCRQ, ACAD9, ATP5MC2, ATP5MC3, COQ10A, COQ10B, COX11, COX14, COX16, COX18, COX20, GLO1, GSTZ1, HAGH, DMAC2L, ECSIT, LDHAL6A, LDHAL6B, ME1, LDHC, LRPPRC, MPC1, MPC2, NDUFAF1, NDUFAF2, NDUFAF3, NDUFAF4, NDUFAF5, NDUFAF6, NDUFAF7, NDUFB11, NUBPL, PPARD, RXRA, SCO1, SLC25A14, SLC25A27, SURF1, TACO1, TIMMDC1, TMEM126B, TRAP1, UQCR10, VDAC1* |
| **Oxidative Phoshorylation**  **(n = 124)** | **GO:0006119** Oxidative phosphorylation  **GO:0090324** Negative regulation of oxidative phosphorylation  **GO:1903862** Positive regulation of oxidative phosphorylation | *ABCD1, ACTN3, AFG1L, ATP5F1A, ATP5F1B, ATP5F1C, ATP5F1D, ATP5F1E, ATP5MC1, ATP5MC2, ATP5MC3, ATP5ME, ATP5MF, ATP5MG, ATP5PB, ATP5PD, ATP5PF, ATP5PO, ATP7A, BID, CCNB1, CDK1, CHCHD10, COA6, COQ9, COX10, COX15, COX4I1, COX4I2, COX5A, COX5B, COX6A1, COX6A2, COX6B1, COX6C, COX7A1, COX7A2, COX7A2L, COX7B, COX7C, COX8A, CYC1, CYCS, DLD, DMAC2L, DNAJC15, DNAJC30, FXN, MECP2, MLXIPL, MSH2, MYOG, NDUFA1, NDUFA10, NDUFA11, NDUFA12, NDUFA13, NDUFA2, NDUFA3, NDUFA4, NDUFA5, NDUFA6, NDUFA7, NDUFA8, NDUFA9, NDUFAB1, NDUFAF1, NDUFB1, NDUFB10, NDUFB11, NDUFB2, NDUFB3, NDUFB4, NDUFB5, NDUFB6, NDUFB7, NDUFB8, NDUFB9, NDUFC1, NDUFC2, NDUFC2-KCTD14, NDUFS1, NDUFS2, NDUFS3, NDUFS4, NDUFS5, NDUFS6, NDUFS7, NDUFS8, NDUFV1, NDUFV2, NDUFV3, NIPSNAP2, PARK7, PDE12, PGK1, PGK2, PINK1, PPIF, RHOA, SDHA, SDHAF2, SDHC, SDHD, SHMT2, SLC25A23, SLC25A33, SNCA, STOML2, SURF1, TAZ, TEFM, UQCC2, UQCC3, UQCR10, UQCR11, UQCRB, UQCRC1, UQCRC2, UQCRFS1, UQCRH, UQCRHL, UQCRQ, VCP* |
| **Mitochondrial DNA**  **(n = 21)** | **GO:0032042** Mitochondrial DNA metabolic process  **GO:0043504** Mitochondrial DNA repair  **GO:0006264** Mitochondrial DNA replication | *CHCHD4, DNA2, FLCN, LIG3, LONP1, MGME1, PARP1, PPARGC1A, SESN2, STOX1, TOP3A, ATG7, DNAJA3, MIR155, POLG, POLG2, PRIMPOL, RRM2B, SSBP1, STOML2, TWNK* |
| **Protein import**  **(n = 19)** | **GO:0030150** Protein import into mitochondrial matrix | *DNAJC15, DNAJC19, DNLZ, GRPEL1, GRPEL2, PAM16, ROMO1, TIMM17A, TIMM17B, TIMM21, TIMM23, TIMM23B, TIMM44, TIMM50, TOMM20, TOMM20L, TOMM40, TOMM40L, TOMM7* |
| **Proton transport**  **(n =15)** | **GO:0033615** mitochondrial proton transporting ATP synthase complex assembly  **GO:0000276** Mitochondrial proton-transporting ATP synthase complex, coupling factor F(o) | *ATP23, ATPAF1, FMC1, OXA1L, TMEM70, ATP5MC1, ATP5MC2, ATP5MC3, ATP5ME, ATP5MF, ATP5MG, ATP5MGL, ATP5PB, ATP5PD, ATP5PF* |

**Supplementary Table 2: Description of the COURAGE-PD consortium.**

| **Cohorts** | **Principal investigators** | **Country *** | **HC** | **PD** |
| --- | --- | --- | --- | --- |
| 1 | Aasly | Norway | 509 | 500 |
| 2 | Annesi | Italy | 94 | 93 |
| 3 | Bardien/Carr | South-Africa | 170 | 280 |
| 4 | Brice/Corvol/Lesage | France | 280 | 779 |
| 5 | Carmine Belin/Ran | Sweden | 628 | 233 |
| 6 | Chartier-Harlin/Muttez | France | 224 | 311 |
| 7 | Deutschlander | Germany | 43 | 257 |
| 8 | Elbaz | France | 1019 | 410 |
| 9 | Farrer | USA | 406 | 284 |
| 10 | Ferreira | Portugal | 54 | 314 |
| 11 | Gasser/Sharma | Germany | 389 | 455 |
| 12 | Duga/Cilia | Italy | 1348 | 1391 |
| 13 | Hadjigeorgiou | Greece | 314 | 267 |
| 14 | Koks/Taba | Estonia | 170 | 216 |
| 15 | Mellick | Australia | 408 | 420 |
| 16 | Puschmann | Sweden | 105 | 55 |
| 17 | Rogaeva/Lang | Canada | 153 | 174 |
| 18 | Stefanis/Simitsi | Greece | 181 | 176 |
| 19 | Valente | Italy | 54 | 213 |
| 20 | Wirdefeldt | Sweden | 171 | 65 |
| 21 | Zimprich | Austria | 184 | 529 |
| **Total** |  |  | 6904 | 7422 |

Country of origin and principal investigators of the 21 European ancestry cohorts composing COURAGE-PD are indicated. GWAS data from a total number of 14326 individuals, including 6904 healthy controls (HC) and 7422 Parkinson’s disease patients (PD), have been made available for this study. * All participants are of European ancestry including those recruited from non-European countries.

**Supplementary Table 3: Summary results of the PRS models performance in the Luxembourg Parkinson’s study and COURAGE-PD cohorts.**

**Luxembourg Parkinson’s Study**

| Gene-set | P-value Threshold | Variance R2 | Adj Variance R2* | FDR P-value | N of clumped SNPs | AUC |
| --- | --- | --- | --- | --- | --- | --- |
| Billingsley I | 0.2627 | 0.012 | 0.0043 | 0.007 | 5371 | 0.55 |
| Billingsley II | 0.1894 | 0.014 | 0.0053 | 0.004 | 28877 | 0.55 |
| MitoCarta 3.0 | 0.0204501 | 0.016 | 0.0058 | 0.003 | 1094 | 0.55 |
| Mitochondrial DNA | 0.1945 | 0.004 | 0.0015 | 0.083 | 408 | 0.52 |
| Mitophagy | 0.0769001 | 0.005 | 0.0021 | 0.053 | 456 | 0.54 |
| Oxidative phosphorylation | 0.0190001 | 0.015 | 0.0058 | 0.003 | 173 | 0.56 |
| TCA cycle | 0.0198001 | 0.006 | 0.0022 | 0.051 | 317 | 0.53 |
| Protein import | 0.2188 | 0.006 | 0.0023 | 0.051 | 345 | 0.52 |
| Proton transport | 0.0948501 | 0.001 | 0.0003 | 0.4 | 108 | 0.5 |

**COURAGE-PD**

| Gene-set | P-value Threshold | Variance R2 | Adj Variance R2* | FDR P-value | N of clumped SNPs | AUC |
| --- | --- | --- | --- | --- | --- | --- |
| Billingsley I | 1 | 0.0091 | 0.00327 | 7.3e-30 | 19602 | 0.52 |
| Billingsley II | 1 | 0.0529 | 0.0195 | 8.5e-153 | 130679 | 0.55 |
| MitoCarta3.0 | 0.37870 | 0.0265 | 0.00964 | 8.4e-82 | 21299 | 0.54 |
| Mitochondrial DNA | 0.49240 | 0.0020 | 0.0007 | 1.2e-07 | 1136 | 0.51 |
| Mitophagy | 0.42580 | 0.0039 | 0.00140 | 7.4e-14 | 2586 | 0.52 |
| Oxidative phosphorylation | 0.07335 | 0.0102 | 0.00366 | 3.4e-33 | 618 | 0.56 |
| TCA_cycle | 1 | 0.0085 | 0.00307 | 3.0e-28 | 14677 | 0.53 |
| Protein import | 0.18590 | 0.0017 | 0.0006 | 7.0e-07 | 274 | 0.51 |
| Proton transport | 0.22530 | 0.001 | 0.0005 | 1.9e-06 | 265 | 0.50 |

Phenotypic variance (R2 score), R2 adjusted for PD prevalence of 0.005*, FDR-adjusted *p* values and predictive accuracy (AUC) of the PRS models are indicated.

**Supplementary Table 4: Primary skin fibroblasts from iPD patients and HC used in functional experiments.**

| **Status** | **Group** | **Fibroblast ID** | **Gender** | **Age at onset (years)** | **Age at biopsy (years)** |
| --- | --- | --- | --- | --- | --- |
| **HC** | **LOW_OXPHOS** | #18486 | M | - | 68 |
|  |  | #18490 | F | - | 62 |
|  |  | #18617 | M | - | 76.5 |
|  |  | #26871 | F | - | 59 |
|  | **HIGH_OXPHOS** | #10365 | M | - | 62.5 |
|  |  | #11020 | M | - | 71 |
|  |  | #15818 | M | - | 72 |
|  |  | #37562 | M | - | 45 |
| **iPD** | **LOW_OXPHOS** | #17162 | M | 66 | 68 |
|  |  | #18250 | M | 57 | 63 |
|  |  | #18263 | M | 52 | 52 |
|  |  | #21990 | M | 70 | 71 |
|  | **HIGH_OXPHOS** | #11043 | M | 68 | 71 |
|  |  | #12251 | M | 50 | 50 |
|  |  | #15092 | M | 66 | 67 |
|  |  | #15809 | F | 40 | 43 |

List of fibroblasts lines established from the Luxembourg Parkinson’s Study participants (both iPD patients and HC) with the highest (HIGH_OXPHOS) or lowest (LOW_OXPHOS) *OXPHOS*-PRSs (10th vs. 90th percentile, n=4 for each group). ID number, gender, age at disease onset and age at the time of skin biopsy are indicated.

**References**

1. Antony PMA, Kondratyeva O, Mommaerts K, Ostaszewski M, Sokolowska K, Baumuratov AS, et al. Fibroblast mitochondria in idiopathic Parkinson’s disease display morphological changes and enhanced resistance to depolarization. Sci Rep. 2020;10:1569.

2. Wasner K, Smajic S, Ghelfi J, Delcambre S, Prada‐Medina CA, Knappe E, et al. Parkin Deficiency Impairs Mitochondrial DNA Dynamics and Propagates Inflammation. Movement Disorders. 2022;mds.29025.

3. Rygiel KA, Grady JP, Taylor RW, Tuppen HAL, Turnbull DM. Triplex real-time PCR–an improved method to detect a wide spectrum of mitochondrial DNA deletions in single cells. Sci Rep. 2015;5:9906.

4. He L. Detection and quantification of mitochondrial DNA deletions in individual cells by real-time PCR. Nucleic Acids Research. 2002;30:68e–68.

5. Billingsley KJ, Barbosa IA, Bandrés-Ciga S, Quinn JP, Bubb VJ, Deshpande C, et al. Mitochondria function associated genes contribute to Parkinson’s Disease risk and later age at onset. npj Parkinsons Dis. 2019;5:8.
